# Transdiagnostic Mental-Health Burden Shapes Cognition and Brain Structure in Adolescents: A Longitudinal Study

**DOI:** 10.64898/2026.06.28.26356781

**Authors:** Pritha Sen, Franziska Knolle

## Abstract

Adolescence is a period of rapid neurodevelopment during which psychiatric symptoms may emerge, yet symptom-specific markers show inconsistent associations with cognition and brain structure and can rarely be generalised longitudinally. Using data from the ABCD Study, we derived a transdiagnostic mental-health burden measure that integrates multiple symptom domains and examined its cognitive and structural brain correlates in early adolescence longitudinally. Adolescents with higher burden showed consistently lower performance in vocabulary, memory, and processing-speed, alongside widespread reductions in whole-brain, cortical, and white-matter volumes at baseline and after 2 years. These effects were strongest in a subgroup with persistent high burden and replicated in cross-sectional analyses. After 4 years, mental-health differences remained robust, although brain-behaviour associations weakened, likely reflecting developmental reorganisation and reduced sample size. Our study demonstrates that global mental-health burden provides a scalable, developmentally appropriate marker of early psychiatric vulnerability that overcomes limitations of symptom-specific approaches.

## 1. Introduction

Adolescence represents a critical period of psychological and neural development, when the brain undergoes extensive structural and functional reorganisation^1,2^, including synaptic pruning of cortices, myelination, and refinement of large-scale connectivity networks^1,3,4^. These maturational processes coincide with the emergence of many psychiatric symptoms and disorders^5,6^. Therefore, this period has been repeatedly identified as a sensitive window for the onset of internalising and externalising symptoms^7–9^, as well as for subclinical experiences such as anxiety, impulsivity, mood instability, and psychosis-like phenomena^10–17^.

Numerous studies have examined individual symptom domains in adolescence, yet their findings remain highly heterogeneous and are often difficult to generalise. Recent work on persistent distressing psychotic-like experiences (PLEs) has identified cognitive disadvantages and global morphometric alterations^18,19^. Meanwhile, studies of internalising and externalising symptoms report domain-specific cognitive profiles^20^, while longitudinal work has linked alterations in cortical development to emerging vulnerability for psychosis, depression and anxiety^21^. These cognitive and neurobiological associations nevertheless vary across cohorts and symptom domains, making it difficult to determine whether observed effects reflect stable vulnerability or transient developmental variation in this period, which is characterised by rapid changes. Although, symptom-focused approaches provide valuable insights into mechanisms, their ability to identify adolescents at broader risk remains limited. Furthermore, it is unclear whether observed alterations represent disorder-specific pathways or shared underlying liabilities.

These developments have motivated a broader conceptual shift toward transdiagnostic and dimensional perspectives^22,23^, which recognise that adolescent symptoms frequently co-occur and share common variance^24,25^. Our previous Adolescent Brain Cognitive Development (ABCD) study on reward anticipation demonstrated that adolescents selected for elevated PLEs and associated distress, also showed elevated depressive, bipolar, anxiety, and suicidality symptoms^26^, suggesting that apparently specific risk markers may reflect broader burden. Furthermore, data-driven approaches, including latent profile analysis^27,28^, structural equation modelling^29^, and clustering^30,31^ capture these shared patterns. However, findings across clustering studies remain heterogeneous, with limited longitudinal validation^30^. This is particularly relevant during early adolescence when symptom expression is fluid, and diagnostic boundaries are still emerging. Several investigations link symptoms to cognitive^19,32–35^ or neuroimaging^19,33,36,37^ alterations, though these analyses are typically cross-sectional^19,32,34–37^, leaving it unclear whether observed differences reflect transient fluctuations or persistent vulnerability. Longitudinal assessments linking mental-health, cognition and neuroanatomy remain comparatively limited, especially in large population-based cohorts. As a result, current approaches may fall short of identifying adolescents who do not yet meet diagnostic thresholds, but who already accumulate elevated transdiagnostic burden and may benefit most from early intervention. From a preventative and clinical perspective, the challenge is therefore not to refine diagnoses at this stage, but to identify youth at an elevated risk before disorders consolidate.

To address this challenge, large longitudinal cohorts such as ABCD provide a unique opportunity to characterise emerging mental-health vulnerability within a developmental framework that integrates symptom burden, cognition, and neuroanatomy over time. Rather than focusing on symptom domains or diagnostic categories in this study, we derived a data-driven transdiagnostic measure of mental-health burden that captures shared liability across multiple symptom dimensions. We identified four latent groups of mental-health burden (very low, low, moderate, high) and examined their associations with cognitive performance and brain structure over five timepoints. By using repeated multimodal assessments during a period when psychopathology remains highly dynamic, this approach enables us to determine whether broad mental-health vulnerability is associated with persistent cognitive and brain differences before diagnostic syndromes become established.

We focused on participants who showed persistent mental-health burden profiles over multiple years to characterize the behavioural and neural correlates of sustained burden and complemented this analysis with cross-sectional assessments using all available data. We hypothesised that adolescents with higher mental-health burden would exhibit lower cognitive performance, and altered brain structure, including differences in cortical thickness and volume, consistent with prior evidence linking psychiatric vulnerability to neurodevelopmental alterations^18,19,38,39^. Furthermore, we expected these associations to be consistent across years, suggesting that sustained mental-health burden reflects a stable vulnerability phenotype with measurable cognitive and neural correlates.

## 2. Methods

### 2.1. Data Descriptions

#### 2.1.1. Participants

Participants were included from the ongoing large-scale longitudinal study, Adolescent Brain and Cognitive Development (ABCD) (release 4.0; https://abcdstudy.org) (11,876 subjects). We included data of 5 time-points: baseline (N=11,868, Age range=9–10 years), 1-year follow-up (N=11,220, Age range=10–11 years), 2-year follow-up (N=10,973, Age range=11–12 years), 3-year follow-up (N=10,336, Age range=12–13 years), and a portion of the 4-year follow-up (N=4,754, Age range=13–14 years), since the complete 4-year follow-up data was unavailable.

#### 2.1.2. Mental-health measures

All youth reported mental-health questionnaires available for each time-point were included in the analysis. Some of these variables were derived from youth-reported K-SADS symptom modules. To maintain a dimensional and single-informant framework, categorical diagnostic variables and parent-reported measures were excluded from the present analyses. The mental-health measures included a broad range of symptom domains such as internalising and externalising symptoms, attentional difficulties, PLEs and associated distress, impulsivity, peer victimization and aggression, life stressors, behavioural inhibition and approach tendencies. These variables encompassed depression, anxiety, bipolar symptoms, sleep problems, suicidality, and impulsivity. Refer to Supplementary Table 1 for further details.

#### 2.1.3. Cognitive measures

Cognitive functioning was assessed using the National Institutes of Health Toolbox Cognition Battery (NIHTB-CB), which includes standardized neuropsychological tests measuring language, executive functioning, working memory, processing speed, and reading ability. Specifically, we analysed scores from the Picture Vocabulary, Flanker Inhibitory Control and Attention, List Sorting Working Memory, Dimensional Change Card Sorting, Pattern Comparison Processing Speed, Picture Sequence Memory, and Oral Reading Recognition tasks. For more details, refer to Weintraub et al., 2013^40^. All scores were used for the analysis.

#### 2.1.4. Structural MRI Measures

Structural MRI data were obtained from T1-weighted scans acquired on 3T scanners (Siemens, GE, or Philips) and processed through the ABCD study’s standardized pipeline using FreeSurfer v5.3.0. From these outputs, we extracted global neuroanatomical measures including mean cortical thickness, total cortical volume, total surface area, and total subcortical volume.

### 2.2. Preprocessing of Mental-Health Measures

We extracted mental-health questionnaire data from the ABCD Study at five timepoints: baseline (n=11,868) and 1- (n=11,220), 2- (n=10,973), 3- (n=10,336), and 4-year (n=4,754) follow-ups. Variables with very low variance, or high number of missing values were excluded (for details, see Section 2.3.1).

For each timepoint, variables with missing data in more than 10% of participants were excluded from the analysis. This procedure eliminated scores for eating disorder and cyber bullying. The remaining missing values were handled using multiple imputation with chained equations (MICE), using predictive mean matching (5 imputations, 5 iterations). Multiple imputations were generated to ensure stable convergence of the imputation model and to avoid relying on a single stochastic draw. As the resulting imputed datasets were nearly identical in this large sample, the first imputed dataset was used for subsequent analyses.

All variables were then rounded to the nearest whole number and truncated at zero to avoid negative values. This helped reduce small artifacts introduced during imputation and ensured consistency across variables for dimensionality reduction.

### 2.3. Mental-Health Burden Score Derivation and Grouping

To capture transdiagnostic mental-health burden across timepoints, we implemented a principal component analysis (PCA) approach to derive a latent mental-health burden score per individual. An alternative approach using z-scored summation of symptoms was used to replicate the PCA-results and is described in the Supplementary Materials Section 2.

#### 2.3.1. PCA approach

To derive a continuous index of transdiagnostic mental-health burden, we applied PCA to the mental-health variables available at each timepoint (baseline to year 4) separately. Only variables with sufficient variability (standard deviation ≥ 0.05) and less than 95% zero values were retained for inclusion. All variables were standardized (z-scored) prior to PCA. The first principal component (PC1), capturing the largest shared variance across domains, was retained as the mental-health burden score.

PC1 explained 24.0% of the variance at baseline, 32.9% at year 1, 24.1% at year 2, 29.7% at year 3, and 24.0% at year 4. Although PC1 explained modest proportion of total variance (24–33%), this is expected in heterogeneous population-based psychiatric datasets comprising multiple partially distinct symptom domains. In such settings, variance is distributed across both general and domain-specific, such that a single component is not expected to account for the majority of variance. Because the primary aim was to quantify broad transdiagnostic burden rather than distinguish specific symptom dimensions, we focused on PC1, which captures variance shared across mental-health domains. Higher-order PCs capture more domain-specific symptom profiles (e.g., dimensions that differentiated particular symptom clusters) rather than the shared variance across domains that was the focus of this study. Complementary analyses using the first three PCs are reported in the Supplementary Material. Importantly, an alternative burden metric based on standardized symptom scores yielded comparable findings across analyses, indicating that findings were not dependent on the PCA-derived measure.

To evaluate whether PC1 explained more variance than expected by chance, we conducted a permutation test (n=5000) in which each variable was independently shuffled and PCA recomputed. The proportion of variance explained by PC1 in the real data was compared to the null distribution. This confirmed that the observed PC1 variance exceeded chance (p<0.001).

To identify distinct participant subgroups, we applied Gaussian Mixture Modeling (GMM) to the mental-health burden score using the mclust package in R^41^. The model selected the optimal number of components (k=4) based on the Bayesian Information Criterion (BIC). Groups were labelled post hoc as *Very Low, Low, Moderate,* and *High Mental-Health Burden* based on their score distributions.

To validate the interpretability of these data-driven groups, we computed group-wise means and 95% confidence intervals for individual symptom scores. Results were visualized using forest plots ordered by effect size, confirming that the derived groups differed meaningfully across multiple symptom domains.

#### 2.3.2. Distributional Comparison of Composite vs. Individual Scores

To evaluate the utility of this transdiagnostic mental-health burden score relative to individual symptom variables, we visualized the score distributions using ridge plots across derived mental-health burden groups. For each year (baseline to 4-year follow-up), we identified the six mental-health variables that showed the largest absolute mean difference between the High and Very Low groups.

#### 2.3.3. Final classification based on group stability

To obtain the final mental-health burden groups, we identified participants who showed persistent or “stable” group membership across 1-, 2- and 3-year follow-ups, meaning that individuals were retained if they were assigned to the same mental-health burden group at each of these three follow-up timepoints. Baseline was excluded from this criterion, as participants were only 9–10 years old, and psychiatric disorder-like symptom presentations in younger children are not always maladaptive. Many behaviours that resemble psychiatric symptoms in adolescents, such as transient social withdrawal, mood fluctuations, or even imaginary friends and psychotic-like experiences (PLEs), may reflect normative aspects of development^42–45^. For instance, imaginative play and vivid fantasy are common learning mechanisms in childhood^46,47^, and some degree of social or emotional liability is expected as adolescents navigate new cognitive and interpersonal demands^48,49^. Symptom patterns tend to become more stable and clinically meaningful as children transition into early adolescence, with many psychiatric disorders first emerging between ages 11 and 14^5,6^. Defining stability beginning after baseline therefore allowed us to focus on sustained patterns of mental-health burden during a developmental window in which emerging vulnerability is more likely to reflect clinically informative trajectories rather than age-typical variation. We also excluded 4-year follow-up data when deriving the mental-health burden groups due to substantial missing data.

Participants with stable scores were then used to define the final mental-health burden groups in all subsequent analyses examining associations with cognitive performance and brain structure.

Although baseline was not used for stability grouping, it is a very important control timepoint to show whether potential neural and behavioural differences are already present earlier. Therefore, to examine whether cognitive and neural differences associated with stable mental-health burden were also detectable outside the years used to define group stability, we used both baseline and 4-year follow-up as control timepoints as described below in Section 2.6.

### 2.4. Group Differences in Cognitive Scores at Year 2 Based on Mental-Health Burden Groups

To examine group differences in cognitive performance, we extracted scores from the NIH Toolbox Cognition Battery and restricted analyses to participants with stable mental-health burden group membership.

We first assessed the number of non-missing observations for each cognitive variable. Variables with insufficient data (missing in 99.3% of participants) were excluded from further analysis. Thus, List Sorting Working Memory and Dimensional Change Card Sorting from Year 2 were dropped.

Group-wise differences were visualized using forest plots showing means and 95% confidence intervals for each cognitive variable, stratified by mental-health burden group (Very Low, Low, Moderate, High). To test statistical significance, we conducted one-way ANOVAs for each cognitive variable using the mental-health burden group as a between-subjects factor. Effect sizes were quantified using eta-squared (η²), and Bonferroni correction was applied to account for multiple comparisons.

### 2.5. Group Differences in Structural Imaging Measures at Year 2 Based on Mental-Health Burden Groups

We also examined group differences in global structural brain measures including cortical thickness, cortical volume, surface area, and subcortical volumes using FreeSurfer-derived imaging parameters from the ABCD dataset.

Left and right hemisphere measures were summed to derive total values (e.g., total cerebral white matter volume=LH + RH). Structural measures were then scaled (z-scored) prior to plotting and statistical testing. To maintain interpretability, we selected eight key structural metrics, consistent with past research^50,51^: whole brain volume, surface area, subcortical grey matter volume, lateral ventricle volume, intracranial volume, cortical volume, cortical thickness, and cerebral white matter volume.

Forest plots were generated for each timepoint, displaying mean differences and 95% confidence intervals across the four mental-health burden groups. One-way ANOVAs were conducted for each variable, followed by Bonferroni corrections for multiple comparisons. Effect sizes were computed using η².

### 2.6. Baseline and Year 4 as a Control Timepoint for Mental-Health Burden Groups

We projected our four mental-health burden groups (Very Low, Low, Moderate, High), originally defined using stable participants across years 1 to 3, onto baseline and year 4 data. For all participants with baseline and available 4-year follow-up data, we compared group-level differences in mental-health variables, cognitive scores, and structural brain measures. These analyses included forest plots to visualize group-wise means and 95% confidence intervals for each domain, followed by one-way ANOVAs to test for statistical differences between mental-health burden groups in each outcome, with Bonferroni correction applied across variables.

These analyses were intended to evaluate how well the predefined mental-health burden groups generalize to a future timepoint not used in model derivation, and whether group-level patterns in mental-health, cognition, and neuroanatomy remain consistent over time.

As a complementary validation approach, we also implemented a z-score-based method to compute mental-health burden scores at each timepoint. This method, along with corresponding GMM and follow-up analyses, is detailed in the Supplementary Materials (Section 2). Results using this approach were consistent with the PCA-based grouping, reinforcing the robustness of our findings.

All cognitive and structural differences throughout were computed including age, sex and site ID as covariates.

### 2.7. Group-wise Analyses Based on Individual-Year Mental-Health Burden Groupings

In addition to analyses using stable participants, we performed complementary analyses based on mental-health burden groups derived separately at each timepoint, to show the validity and clinical relevance of this approach in the absence of longitudinal data. Specifically, mental-health burden scores were computed using the PCA-based approach at each timepoint, and participants were assigned to four mental-health burden groups (Very Low, Low, Moderate, High) using GMM applied independently at each year.

For each timepoint, we examined group-level differences in cognitive performance and structural brain measures using ANOVAs, followed by Bonferroni correction for multiple comparisons. This allowed us to assess whether the associations between mental-health burden and cognitive or structural indices persisted when not constrained by longitudinal group stability.

### 2.8 General Procedures

All statistical analyses were conducted using the R Statistical Software (version 4.4.2)^52^. Variable standardization was performed using the base R function scales (version 1.4.0)^53^, and missing data were handled using multiple imputation via the mice package (version 3.17.0)^54^. Visualization of group distributions, forest plots, and transitions over time was carried out using ggplot2 (version 3.5.2)^55^, ggalluvial (version 0.12.5)^56^, cowplot^57^ and ggridges (version 0.5.6)^58^. The progress package (version 1.2.3)^59^ was used to monitor computational steps during permutation testing. To identify latent mental-health burden groups, we applied GMM using the mclust package (version 6.1.1)^41^. Dimensionality reduction via PCA was implemented using base R functions. Group comparisons were conducted using rstatix (version 0.7.2)^60^, which provided functions for ANOVA and chi-square testing, and effectsize (version 1.0.0)^61^ was used to compute standardized effect sizes and confidence intervals. Given the large sample size (n ≈ 11,000), parametric analyses were considered robust to deviations from normality.

## 3. Results

### 3.1. Mental-Health Variables Differences

We first examined whether the data-driven mental-health burden groups showed meaningful separation on the underlying mental-health variables that were used to derive them. Forest plots revealed clear and consistent stratification across mental-health burden groups (Very Low, Low, Moderate, High) at each timepoint from baseline to 3-year follow-up. Across all panels in Figure 1 (A–E), participants in the High mental-health burden group displayed the highest scores across nearly all symptom dimensions, particularly for internalizing symptoms such as depression, anxiety, suicidality, distress caused due to PLEs, and bipolar disorder symptoms. In contrast, the Very Low group consistently showed the lowest levels of psychopathology.

**Figure 1.**
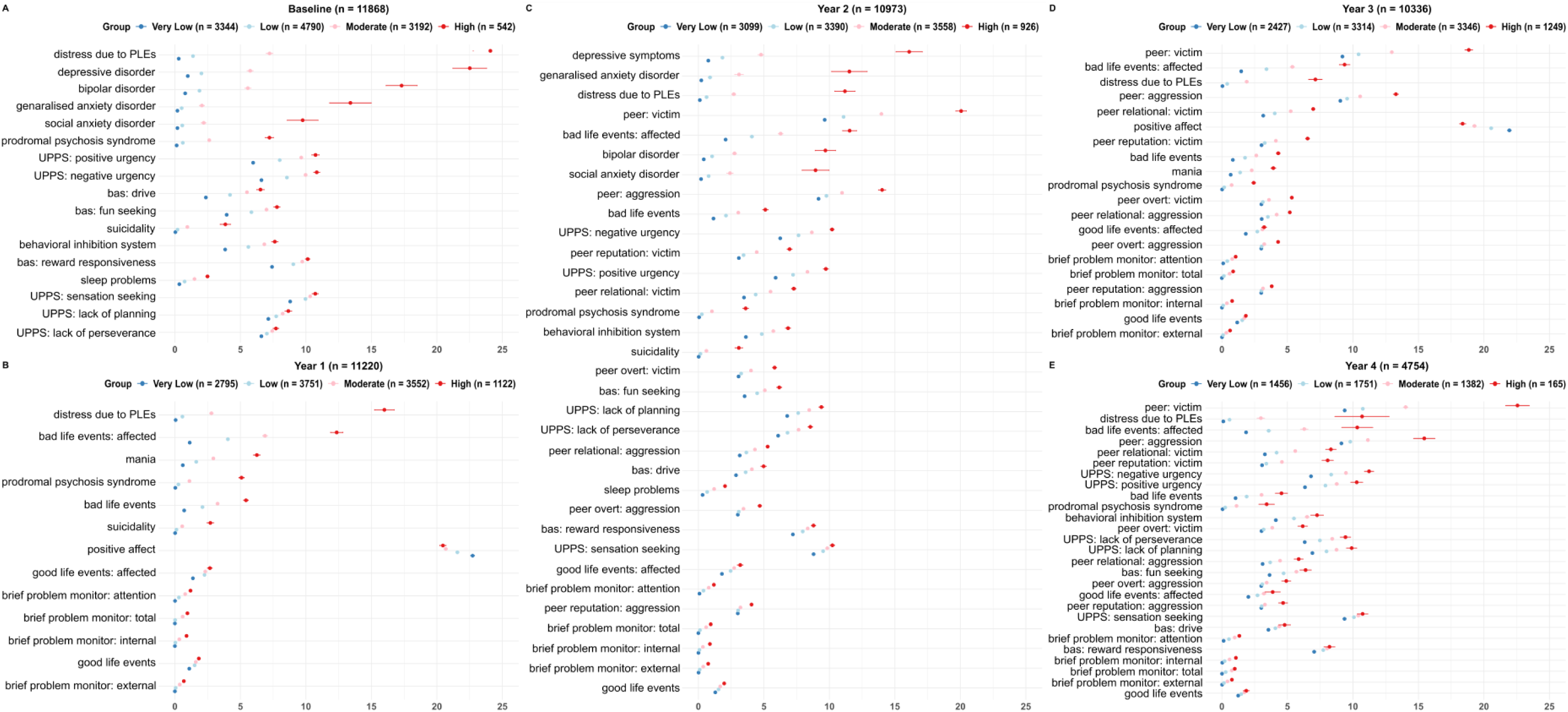
Forest plots showing group-wise comparisons of mental-health scores across timepoints. Note: Participants were grouped into four mental-health burden categories (Very Low, Low, Moderate, High) based on Gaussian Mixture Modelling applied to the first principal component of mental-health scores at each timepoint. Points reflect group means and 95% confidence intervals. Variables within each panel are ordered by effect size for the High vs. Very Low group comparison. Clear group separation is observed across all timepoints. Abbreviations: PLEs = psychotic-like experiences; UPPS = Urgency, Premeditation, Perseverance, Sensation Seeking, Positive Urgency, Impulsive Behaviour; bas = behavioural approach system. Please refer to Supplementary Table 1 for more details on the scores.

At baseline (Figure 1A), the clearest separation between mental-health burden groups was observed for distress caused due to PLEs (distress), depression, prodromal psychosis syndrome (pps), social and generalized anxiety disorder, and bipolar disorder scores. These variables showed a graded increase from the Very Low to High group, with the High group displaying particularly elevated scores. The remaining variables also contributed to the separation, albeit to a slightly lesser extent.

These group differences remained consistent in Year 1 (Figure 1B), with distress, pps, and bad life event impact continuing to show strong differentiation across groups. Variables such as mania and suicidality were also notably elevated in the High group during this timepoint.

In Year 2 (Figure 1C), the separation persisted and expanded across a broader set of variables, including depression, generalized anxiety disorder, distress caused due to PLEs, peer victimization, bad life event impact and bipolar disorder. The High group consistently exhibited higher levels across these dimensions compared to other groups.

By Year 3 (Figure 1D), peer victimization, relational victimization, peer aggression and positive affect emerged as particularly distinguishing features of the High group. While symptoms such as bad life event impact and distress caused due to PLEs remained elevated, peer-related difficulties became increasingly prominent contributors to mental-health burden differentiation at this later stage.

By Year 4 (Figure 1E), the High burden group continued to report elevated scores with maximum differences in distress caused due to PLEs, affected bad life events, peer victimization and related subscales and peer aggression compared to lower burden groups.

These temporal patterns were further supported by the ridge density plots (Figure 2 A–E), which visualize the top six variables contributing to group separation at each year. Importantly, distress caused due to PLEs and bad life event impact consistently appeared across years as top contributors, while peer-related variables became more dominant in later years.

**Figure 2.**
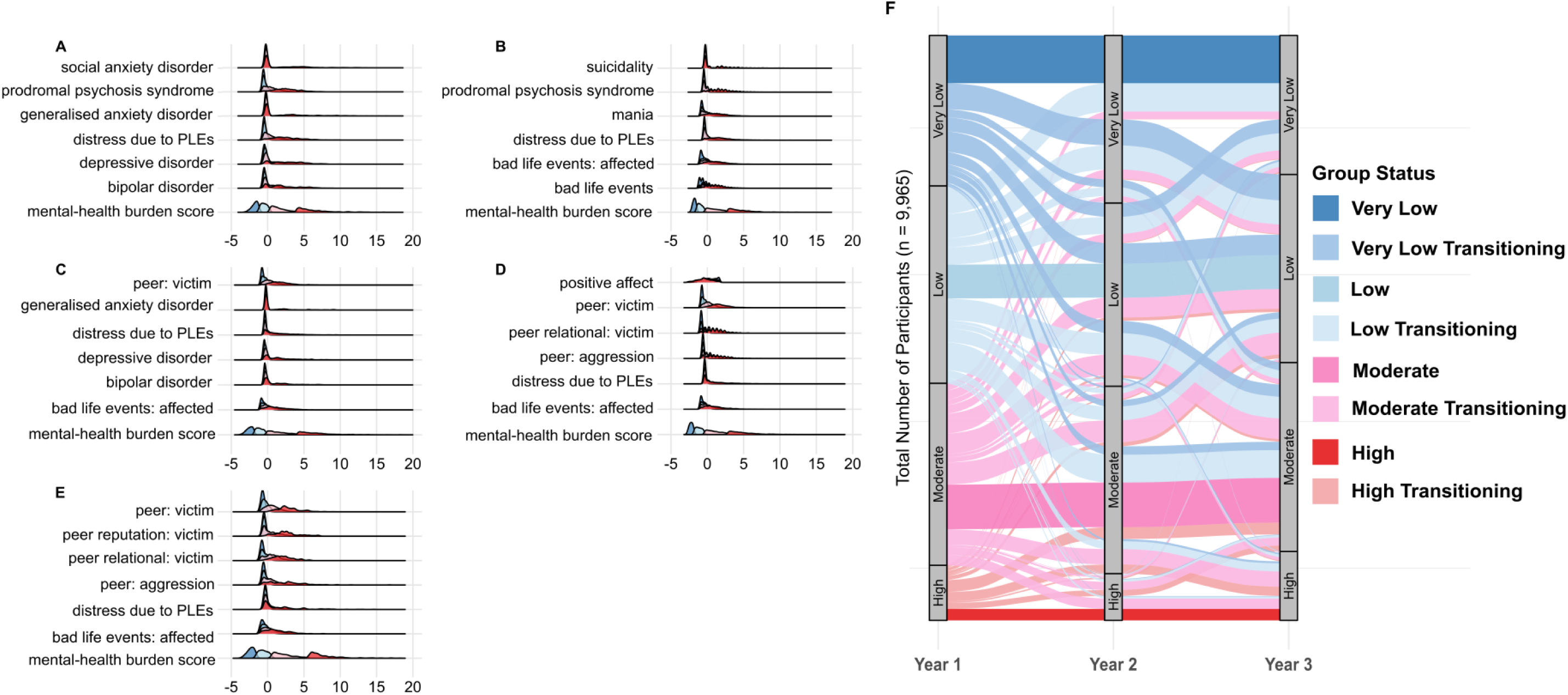
Distribution plots showing mental-health burden group separation, and sankey diagram showing mental-health burden group transitions over time. Note: (A) to (E) For each year (baseline to Year 4), the six mental-health scores shown are those with the largest absolute mean difference between the High and Very Low groups; x-axis displays the scores of the respective symptoms. Abbreviation: PLEs = psychotic-like experiences. (F) The plot displays transitions in mental-health burden group membership from 1- to 3-year follow-up among participants with complete data across all four timepoints (*n*=9,965). Groups were defined using PCA and Gaussian Mixture Modelling at each timepoint. Participants who remained in the same group from 1- to 3-year follow-up are labelled by their stable group (e.g., Low) and indicated in darker colours, while those who changed groups are labelled as “Transitioning” from their 1-year follow-up classification (e.g., Low Transitioning) and indicated in lighter colours. Stream widths represent the number of individuals transitioning between groups.

Notably, effect sizes remained substantial and stable across years (please see Table 1), underscoring the robustness of the PCA-derived mental-health burden classification in capturing meaningful and persistent symptom differences.

**Table 1.** Group differences in mental-health variables based on individual year mental-health burden groups. Note: Due to their size, Tables 1–3 and Supplementary Tables 1–4 are provided as a separate supplementary Excel file.

To support the rationale for deriving a transdiagnostic mental-health burden score, we visualized the distribution of mental-health variables across mental-health burden groups from Baseline to 4-year follow-up. For each timepoint, we selected the top six individual symptom variables based on effect size differences between mental-health burden groups. Group-wise density plots were generated for each of these variables alongside the composite mental-health burden score. As shown in Figure 2(A–E), individual variables (e.g., distress caused due to PLEs, bad life events) showed considerable distributional overlap between groups. In contrast, the composite score demonstrated clearer separation across groups (Very Low to High), highlighting its improved sensitivity and interpretability. These plots reinforce the benefit of aggregating across multiple domains to derive a more robust and continuous index of transdiagnostic mental-health burden.

### 3.2. Mental-Health Burden Group Transitions from 1- to 3-year follow-up

To explore how participants’ mental-health burden group assignments evolved over time, we visualized transition patterns across 1-, 2- and 3-year follow-ups using a Sankey diagram (Figure 2F). This analysis included all participants (n=9,965) who had complete data across these three timepoints.

The width of each stream in the diagram represents the number of individuals transitioning from one group to another. Group memberships were defined separately at each timepoint using the PCA + GMM pipeline. To highlight patterns of longitudinal stability, participants whose group assignments remained consistent from Year 1 through Year 3 were labelled by their stable group. In contrast, participants whose group assignments fluctuated across years were labelled based on their Year 1 group and marked as “Transitioning”.

The visualization revealed that while many participants retained their original group classification over time, there was also notable group mobility. Overall, 23.6% of participants maintained the same group membership from Year 1 to Year 3. Stability varied by group, with the Very Low group showing the highest retention (54.4%), followed by the Moderate (47.9%), Low (38.2%), and High groups (34.7%). Transitions in either direction occurred predominantly between adjacent groups, particularly between the Low and Moderate burden levels, whereas direct shifts between the Very Low and High groups were rare (<1%). These findings suggest moderate longitudinal continuity of mental-health burden, with most changes occurring between neighbouring burden categories rather than across the full burden spectrum.

These dynamic patterns underscore the importance of tracking intra-individual variability in mental-health burden over time and validating the presence of both stable and fluctuating trajectories within the PCA-based grouping framework.

Participants who showed stable group membership across 1-, 2- and 3-year follow-ups defined the final mental-health burden groups (Very Low: n=818, Low: n=577, Moderate: n=760, High: n=196). Demographic characteristics for stable participants are presented in Table 2. Further details on mental-health variable distributions for stable participants are presented in Table 3.

**Table 2.** Demographic characteristics for stable participants with persistent mental-health burden groups from 1-year follow-up to 3-year follow-up.

**Table 3.** Group differences in mental-health variables for stable participants based on persistent mental-health burden groups from 1-year follow-up to 3-year follow-up.

### 3.3. Cognitive Performance Across Final Mental-Health Burden Groups

We next examined whether PCA-derived mental-health burden groups differed in cognitive performance at Year 2, using NIH Toolbox scores. As shown in Figure 3, group-wise differences were observed across multiple domains, with the High mental-health burden group consistently showing poorer performance.

**Figure 3.**
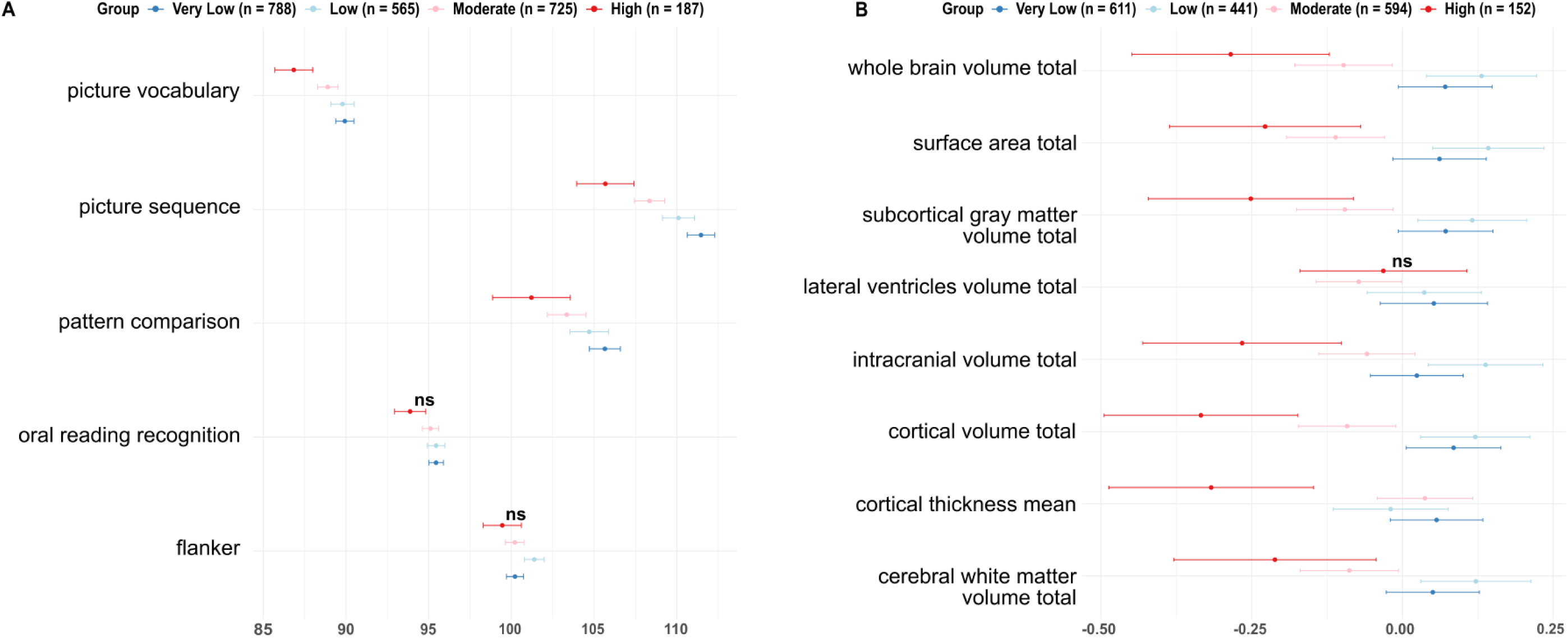
Group differences in cognitive performance and brain structure at baseline and 2-year follow-up, by mental-health burden group based on stable participants across 1-year follow-up through 3-year follow-up Note: (A) Cognitive tasks scores at 2-year follow-up. The High mental-health burden group scored lower across most domains. Group differences were particularly pronounced in Picture Vocabulary, Picture Sequence, and Pattern Comparison tasks. (B) Structural measures 2-year-follow-up time-point. Left and right hemisphere measures were summed to derive total values (e.g., cerebral white matter volume total =LH + RH) and subsequently standardised. Points reflect group means and 95% confidence intervals. ns indicates non-significant group differences (p<0.05) after correcting for multiple comparisons.

At 2-year follow-up (Figure 3A), significant differences emerged across three cognitive domains including picture vocabulary (F(3,2171)=8.8, p<0.001, η²=0.01), picture sequence (F(3,2182)=15.26, p<0.001, η²=0.02), pattern comparison (F(3,1774)=5.05, p=0.009, η²=0.009). No group differences were observed for flanker (F(3,1780)=3.74, p=0.053, η²=0.006) or oral reading recognition (F(3,2166)=3.29, p=0.099, η²=0.005) scores at Year 2.

Overall, these findings indicate that youth with persistently high mental-health burden show lower cognitive performance in domains involving language, memory, and processing speed.

All results are corrected for multiple comparisons using Bonferroni correction (p<0.05).

### 3.4. Structural Imaging Differences Across Final Mental-Health Burden Groups

We then examined whether mental-health burden groups differed in global structural brain measures at the 2-year follow-up. At 2-year follow-up (Figure 3B), we observed significant group differences in nearly all structural features, including whole brain volume (F(3,1749)=13.37, p<0.001, η²=0.02), surface area (F(3,1749)=12.17, p<0.001, η²=0.02), subcortical grey matter volume (F(3,1749)=10.53, p<0.001, η²=0.02), intracranial volume (F(3,1749)=11.52, p<0.001, η²=0.02), cortical volume (F(3,1749)=14.73, p<0.001, η²=0.02), mean cortical thickness (F(3,1749)=6.54, p=0.002, η²=0.01) and cerebral white matter volume (F(3,1749)=8.47, p<0.001, η²=0.01). All measures except lateral ventricle volume (F(3,1749)=2.04, p=0.853, η²=0.003) showed significant group differences after correction for multiple comparisons. All results are corrected for multiple comparisons using Bonferroni correction (p<0.05).

These results suggest that youth with higher mental-health burden show widespread differences in global brain structures.

### 3.5. Results for Baseline and Year 4 as Control Timepoints for Final Mental-Health Burden Groups

To evaluate whether the mental-health burden groupings derived from stable trajectories (Year 1 to Year 3) retained predictive utility at baseline and Year 4, we examined group-level differences in mental-health variables, cognitive and structural outcomes at these time-points, using these stable group assignments. Here, the available MRI and cognitive sample sizes were substantially smaller (60%) than in previous years. Please refer to Table 3 for further details on mental-health score distributions and differences for baseline and 4-year follow-up as control datasets.

We observed widespread and consistent group differences in mental-health variables. All variables showed statistically significant differences across mental-health burden groups after Bonferroni correction (p<0.05), with the exception of UPPS Sensation Seeking at Year 4 (Figure 1A and 1E; see Table 3 for full ANOVA statistics). In contrast, differences in cognitive and structural measures at Year 4 were less consistent than in earlier timepoints, likely influenced by the markedly reduced sample size.

At baseline (Figure 4A), significant group differences emerged in five out of seven cognitive tasks, including picture vocabulary (F(3,2319)=5.25, p=0.009, η²=0.007), picture sequence (F(3,2318)=13.78, p<0.001, η²=0.02), pattern comparison (F(3,2314)=7.07, p<0.001, η²=0.009), list sorting working memory (F(3,2314)=12.44, p<0.001, η²=0.02), and dimensional change card sorting (F(3,2320)=7.39, p<0.001, η²=0.01). In contrast, no statistically significant differences were observed in flanker (F(3,2317)=2.5, p=0.403, η²=0.003) or oral reading recognition F(3,2318)=2.97, p=0.214, η²=0.004) scores at baseline.

**Figure 4.**
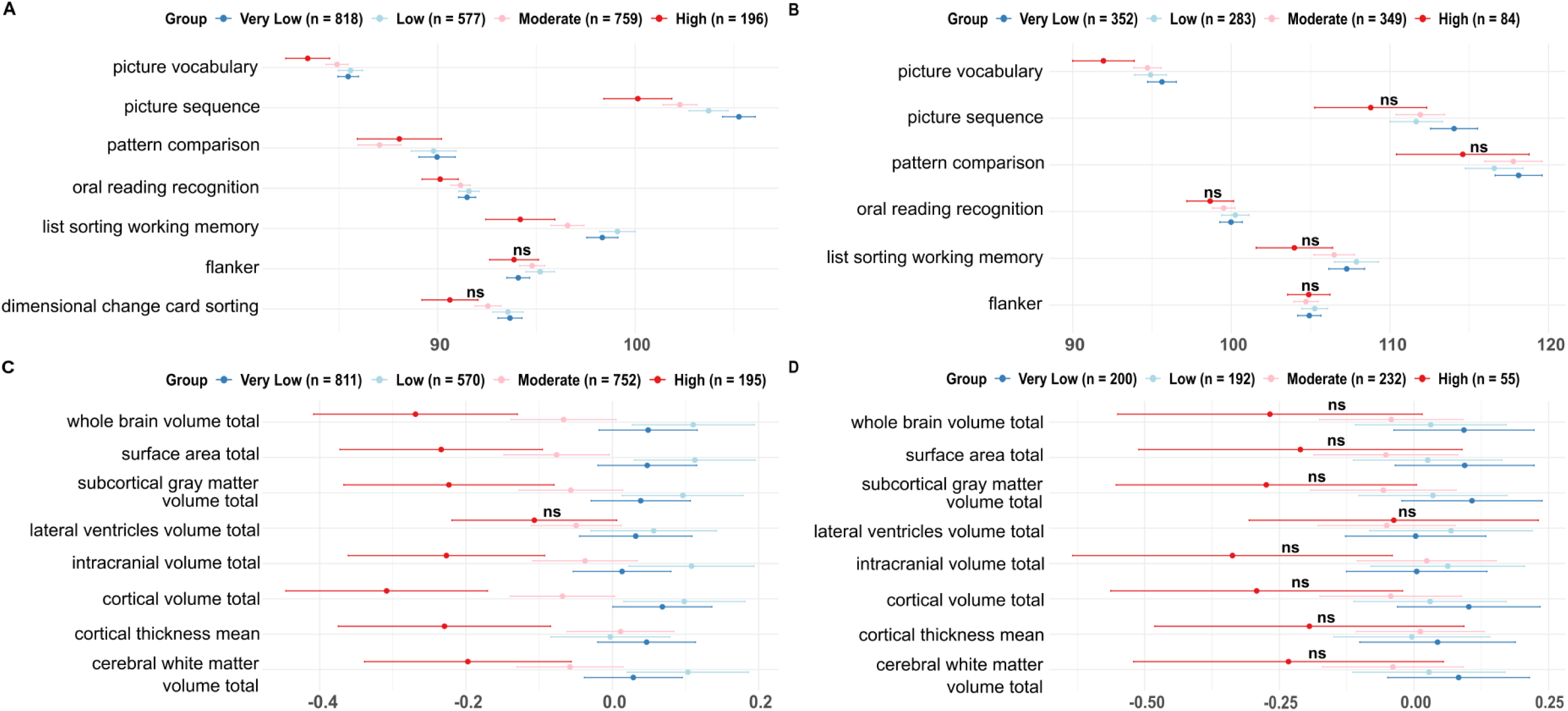
Group differences in cognitive performance and structural measures at control timepoints: baseline and 4-year follow-up, by mental-health burden group based on stable participants across 1-year follow-up through 3-year follow-up. Note: Upper panel – cognitive tasks scores at (A) baseline, (B) 4-year follow-up. Lower panel – structural measures at (C) baseline and (D) 4-year-follow-up time-point. Left and right hemisphere measures were summed to derive total values (e.g., cerebral white matter volume total =LH + RH) and subsequently standardised. Points reflect group means and 95% confidence intervals. ns indicates non-significant group differences (p<0.05) after correcting for multiple comparisons.

Among the NIH Toolbox cognitive measures (Figure 4B) available at Year 4, the Picture Vocabulary test (F(3,1041)=4.49, p=0.023, η²=0.01) continued to show a significant group difference after Bonferroni correction (p<0.05). All other cognitive tests (Flanker (F(3, 732)=0.24, p=1, η²=0.001), List Sorting Working Memory (F(3,1040)=2.78, p=0.24, η²=0.008), Pattern Comparison (F(3,731)=1.02, p=1, η²=0.004), Picture Sequence (F(3,1037)=3.54, p=0.086, η²=0.01) and Oral Reading Recognition (F(3,1035)=1.43, p=1, η²=0.004)) did not differ significantly between groups.

At baseline (Figure 4C), we observed significant group differences in nearly all structural features, including whole brain volume (F(3,2323)=12.3, p<0.001, η²=0.02), surface area (F(3,2323)=11.39, p<0.001, η²=0.01), subcortical grey matter volume (F(3,2323)=8, p<0.001, η²=0.01), intracranial volume (F(3,2323)=9.68, p<0.001, η²=0.001), cortical volume (F(3,2323)=13.86, p<0.001, η²=0.02), mean cortical thickness (F(3,2324)=4.55, p=0.028, η²=0.006) and cerebral white matter volume (F(3,2323)=7.5, p<0.001, η²=0.01). No significant differences were observed in lateral ventricle volume (F(3,2323)=2.34, p=0.569, η²=0.003).

At Year 4, none of the global structural brain measures (Figure 4D) (whole brain volume (F(3,670)=3.4, p=0.139, η²=0.02), surface area (F(3,670)=2.55, p=0.44, η²=0.01), subcortical grey matter volume (F(3,670)=3.64, p=0.101, η²=0.02), lateral ventricle volume (F(3,670)=0.681, p=1, η²=0.002), intracranial volume (F(3, 670)=3.83, p=0.078, η²=0.02), cortical volume (F(3, 670)=3.96, p=0.065, η²=0.02), cortical thickness (F(3,670)=0.88, p=1, η²=0.004) and cerebral white matter volume (F(3,670)=2.34, p=0.578, η²=0.01)) showed significant differences across mental-health burden groups after correction for multiple comparisons.

This suggests that while group differences in brain structure were observable at earlier timepoints (baseline and Year 2), these distinctions may not persist or may be less detectable at later stages, especially given the reduced sample size and partial data availability at Year 4. However, mental-health differences across groups remained robust, suggesting that the mental-health burden grouping continues to capture relevant clinical variation even at later stages.

### 3.6. Group Differences in Cognitive Performance Based on Individual-Year Mental-Health Burden Groups

In contrast to the previous analyses based on stable mental-health burden trajectories, the results below present cross-sectional analyses in which mental-health burden groups are defined independently at each timepoint.

At baseline (Figure 5A), significant group-level differences were observed for all cognitive tasks, including picture vocabulary (F(3,11716)=104.48, p<0.001, η²=0.03), flanker (F(3,11710)=11.17, p<0.001, η²=0.003), picture sequence (F(3,11704)=31.99, p<0.001, η²=0.008), pattern comparison (F(3,11692)=37.7, p<0.001, η²=0.01), list sorting working memory (F(3,11667)=68.63, p<0.001, η²=0.02), dimensional change card sorting (F(3,11711)=53.14, p<0.001, η²=0.01) and oral reading recognition (F(3,11702)=60.06, p<0.001, η²=0.02).

**Figure 5.**
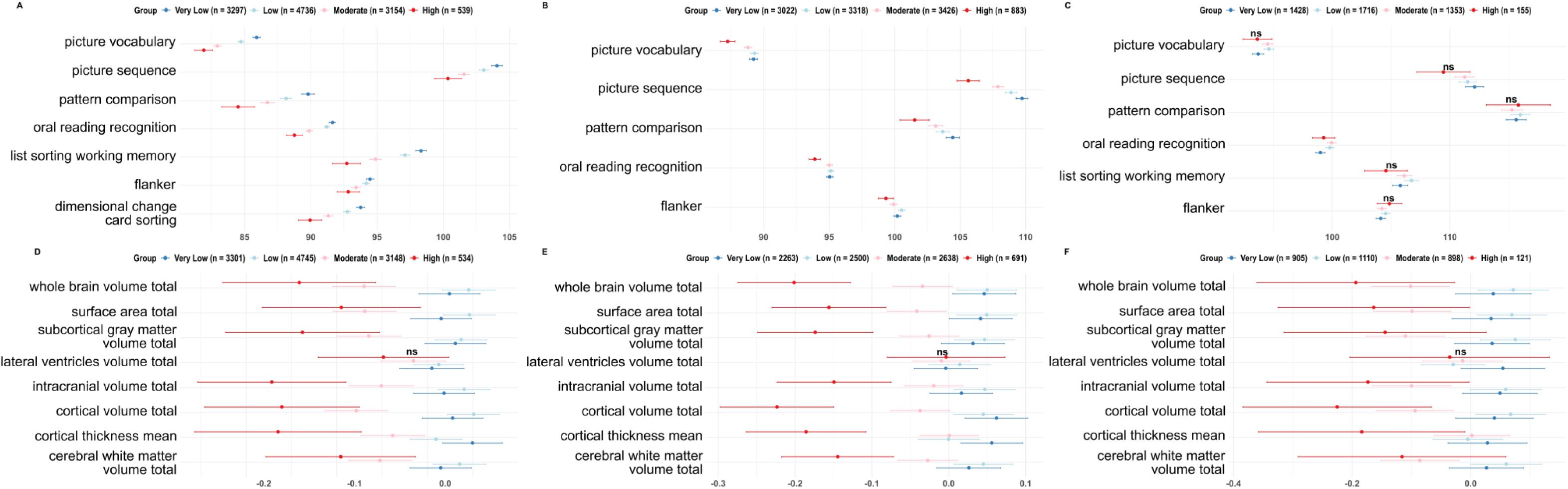
Group differences in cognitive performance and brain structure at baseline, 2-year follow-up and 4-year follow-up, based on individual year mental-health burden groups. Note: Upper panel – cognitive tasks scores at (A) baseline, (B) 2-year follow-up and (C) 4-year follow-up; Lower panel – structural measures at (D) baseline, (E) 2-year follow-up and (F) 4-year follow-up. Left and right hemisphere measures were summed to derive total values (e.g., cerebral white matter volume total =LH + RH) and subsequently standardised. Points reflect group means and 95% confidence intervals. ns indicates non-significant group differences (p<0.05) after correcting for multiple comparisons.

Similarly, at Year 2 (Figure 5B), group-level differences were observed for all cognitive tasks, including picture vocabulary (F(3,9844)=13.53, p<0.001, η²=0.004), flanker (F(3,7926)=5.25, p=0.006, η²=0.002), picture sequence (F(3,9873)=26.55, p<0.001, η²=0.008), pattern comparison (F(3,7888)=8.15, p<0.001, η²=0.003) and oral reading recognition (F(3,9805)=7, p<0.001, η²=0.002).

At Year 4 (Figure 5C), after correcting for multiple comparisons, significant differences were observed only for oral reading recognition (F(3,4532)=5.3, p=0.007, η²=0.003). The other scores, including picture vocabulary (F(3,4562)=3.83, p=0.057, η²=0.002), flanker (F(3,3264)=0.93, p=1, η²=0.0009), picture sequence (F(3,4534)=1.76, p=0.92, η²=0.001), pattern comparison (F(3,3260)=0.35, p=1, η²=0.0003) and list sorting working memory (F(3,4535)=2.66, p=0.281, η²=0.002), remained non-significant.

All results are corrected for multiple comparisons using Bonferroni correction (p<0.05).

Taken together, these results show that mental-health burden groups defined at individual timepoints are associated with widespread cognitive performance differences, particularly at earlier timepoints. At baseline and 2-year follow-up, the High mental-health burden group consistently showed lower scores across multiple cognitive domains, especially in picture vocabulary, working memory, and pattern comparison tasks, aligning with effect size gradients visible in Figure 5A–B. However, by Year 4, most group differences were no longer statistically significant, suggesting a potential convergence of cognitive scores over time or greater inter-individual variability in later adolescence. The only exception was oral reading recognition, which remained significantly different across groups, albeit with small effect size.

### 3.7. Group Differences in Brain Structure Based on Individual-Year Mental-Health Burden Groups

In this section, we examined structural brain differences using the cross-sectional mental-health burden groups defined independently at each timepoint.

Structural MRI measures also revealed significant group-level differences at both timepoints when using year-wise mental-health burden groups. At baseline and year 2, almost all structural measures, except for lateral ventricle volume, showed significant variation across groups, with higher mental-health burden linked to reduced whole-brain, cortical, and white matter volumes, among others. At year 4, all measures except lateral ventricle volume and cortical thickness were significant different across groups.

Baseline (Figure 5D): whole brain volume (F(3,11724)=17.04, p<0.001, η²=0.004), surface area (F(3,11724)=14.04, p<0.001, η²=0.004), subcortical gray matter volume (F(3,11724)=13.9, p<0.001, η²=0.004), lateral ventricle volume (F(3,11724)=0.97, p=1, η²=0.0002), intracranial volume (F(3,11724)=16.45, p<0.001, η²=0.004), cortical volume (F(3,11724)=20.61, p<0.001, η²=0.005), cortical thickness (F(3,11724)=10.26, p<0.001, η²=0.003), cerebral white matter volume (F(3,11724)=8.84, p<0.001, η²=0.002).

Year 2 (Figure 5E): whole brain volume (F(3,8088)=19.1, p<0.001, η²=0.007), surface area (F(3,8088)=14.26, p<0.001, η²=0.005), subcortical grey matter volume (F(3,8088)=12.77, p<0.001, η²=0.005), lateral ventricle volume (F(3,8088)=0.28, p=1, η²=0.0001), intracranial volume (F(3,8088)=10.99, p<0.001, η²=0.004), cortical volume (F(3, 8088)=22.89, p<0.001, η²=0.008), cortical thickness (F(3,8088)=11.37, p<0.001, η²=0.004), cerebral white matter volume (F(3,8088)=9.91, p<0.001, η²=0.004).

Year 4 (Figure 5F): whole brain volume (F(3,3030)=10.06, p<0.001, η²=0.01), surface area (F(3,3030)=8.72, p<0.001, η²=0.007), subcortical grey matter volume (F(3,3030)=9.36, p<0.001, η²=0.009), lateral ventricle volume (F(3,3030)=1.36, p=1, η²=0.001), intracranial volume (F(3,3030)=9.21, p<0.001, η²=0.009), cortical volume (F(3,3030)=9.76, p<0.001, η²=0.01), cortical thickness (F(3,3030)=1.8, p=1, η²=0.002), cerebral white matter volume (F(3,3030)=5.65, p=0.006, η²=0.006).

All results are corrected for multiple comparisons using Bonferroni correction (p<0.05).

The structural differences were similar to the cognitive findings, with the most pronounced and consistent effects observed at baseline and 2-year follow-up. Across both timepoints, individuals in the High mental-health burden group showed lower global brain metrics, as shown in Figure 5D–E. These effects remained statistically robust after correction, with small but consistent effect sizes. Notably, lateral ventricle volume showed no group differences at any timepoint. At Year 4 (Figure 5F), significant group differences persisted across brain volume metrics, but the magnitude of these effects appeared attenuated compared to earlier years.

## 4. Discussion

The present study used a large longitudinal population cohort (ABCD) to identify a data-driven indicator of adolescent mental-health burden and to test its behavioural and neurobiological correlates. Using both stability-based and complementary cross-sectional analyses, we demonstrate that transdiagnostic mental-health burden is consistently and longitudinally associated with differences in cognitive performance and global brain structure during adolescence. In contrast to symptom-specific or purely cross-sectional approaches, our framework captures sustained cross-domain vulnerability and links it to neurodevelopmental variation. The findings therefore extend prior dimensional and transdiagnostic models of youth psychopathology, which have largely focused on single symptom domains or cross-sectional latent profiles^19–21^. By quantifying a broad burden dimension that cuts across traditional diagnostic boundaries, this approach may be particularly useful for identifying adolescents at elevated risk before discrete disorders emerge and diagnostic syndromes consolidate, when preventative intervention strategies targeting general psychological wellbeing and resilience may be most effective.

A key strength of our approach lies in focusing on adolescents who maintained the same mental-health burden profile from Year 1 to Year 3, thereby capturing sustained vulnerability during a developmental window when symptoms begin to consolidate^7,62^. Within this approach, we observed robust associations with cognition and global brain measures at Year 2, while comparable differences were already detectable at baseline before stable profiles were defined. Together, this supports the interpretation that sustained mental-health burden reflects a meaningful liability phenotype and potential pre-diagnostic marker of neurodevelopmental vulnerability^63,64^. This is consistent with prior longitudinal work showing that persistent multi-domain problems substantially increase the likelihood of formal psychiatric diagnosis^62^, reinforcing that those who maintain elevated burden over time represent a distinct higher-risk subgroup. Notably, adolescents who were later classified into stable high-burden groups already showed cognitive and structural differences at baseline, before their stable profiles were defined. This suggests that neurodevelopmental differences associated with transdiagnostic burden begin to emerge prior to symptom consolidation, potentially representing early markers of vulnerability. These associations replicated in cross-sectional analyses using the full sample, further reinforcing the reliability and generalisability of the findings.

The strongest mental-health differences were driven by distressing PLEs, depression, anxiety, bipolar symptoms, suicidality, mania, disturbing life events, and peer adversity, underscoring the multi-domain nature of this vulnerability. However, as not all measures were available at every timepoint, future work with consistent longitudinal measurement will be needed to confirm their relative contributions. With regard to cognition, adolescents with high mental-health burden consistently performed worse across vocabulary, episodic memory, processing speed, and working memory. Prior work focusing on domain-specific symptoms has linked depression to verbal memory and fluency deficits^65^, and ADHD to executive dysfunction^66^. In contrast, the broad impairments observed in our study align with evidence that cross-domain psychopathology burden is associated with broader cognitive deficits than internalising or externalising symptoms alone^35^. Examining brain structure, we found widespread reductions across whole-brain, cortical, and white-matter volumes with higher mental-health burden. This pattern is consistent with prior evidence linking global brain volume to persistently elevated general psychopathology^67^, as well as studies showing that internalising-externalising comorbidity is associated with regional structural alterations beyond the effects of either domain alone^68^. Lateral ventricle volume did not differ between groups, suggesting that structural correlates of cumulative burden are reflected in reductions in brain tissue volume rather than ventricular expansion. This is consistent with evidence that ventricular enlargement is absent in subclinical and high-risk samples^69^ but observed in adolescents and adults with psychotic symptoms^70^. Notably, distinctions were clearest between Very Low/Low groups and Moderate/High groups, with Very Low/Low groups showing relatively similar structural profiles. Future work examining regional or network-specific measures may further elucidate these patterns. Together, these findings position cumulative mental-health burden as a unifying framework capable of integrating previously fragmented domain-specific observations.

At Year 4, group differences in cognition and global brain structure were attenuated in the stability-based analyses, despite substantial differences in mental-health burden between groups. Reduced sample sizes for cognitive and structural data, but also increased pubertal neurodevelopmental variability^2,4^, and evolving symptom trajectories^71^, likely contributed to this attenuation. Importantly, the contemporaneous cross-sectional analyses at Year 4 continued to show strong structural differences, indicating that mental-health burden remains behaviourally and neurobiologically relevant during this developmental period. The attenuated global effects in the longitudinal analysis, should therefore not be interpreted as evidence that vulnerability has disappeared, but rather as a combination of developmental and methodological factors, which future analyses may clarify.

While recent work in the same cohort shows that persistent distressing PLEs predict cognitive decline and brain morphometric changes^18^, our findings suggest that such alterations may not be exclusive to PLEs alone, but may reflect broader psychiatric burden across multiple symptom domains. Importantly, our study showed convergence across both stability-based and cross-sectional analyses and yielded consistent cognitive and structural findings in PCA as well as z-score-based classification approaches (see Supplementary materials for z-score-based grouping results).

Certain limitations should be noted, including reduced sample size for Year 4 assessments and reliance on global brain measures. Future work should incorporate regional or network-level brain measures and evaluate alternative definitions of persistent mental-health burden, such as late-onset increases or convergence into higher-burden groups, capturing additional subtypes of vulnerability.

Early identification of psychiatric vulnerability remains a central goal in adolescent mental-health research^72^, as intervention during this period may substantially alter long-term outcomes. However, early adolescence is characterised by the emergence of a wide array of psychiatric symptoms that frequently flare up across domains and are often diffuse and developmentally unspecific. Although individual symptoms have been linked to cognitive and neuroanatomical alterations, these associations are typically weak, inconsistent, and unstable across development, and may therefore overlook many individuals who are already accumulating meaningful vulnerability. Addressing these challenges, we proposed a transdiagnostic burden framework as a more developmentally appropriate and clinically scalable approach for identifying vulnerable youth. By demonstrating robust cognitive and neuroanatomical alterations associated with sustained burden, replicated across analytic strategies, our results support a burden-oriented assessment strategy. This longitudinal mental-health burden approach may offer a more stable route for early detection and guiding interventions such as cognitive-behavioural therapy, social skills training, or cognitive remediation. Future work should determine the minimal set of symptom domains required to derive a reliable burden measure across cohorts, with the ultimate goal of developing a scalable and transferable framework for early identification and prevention.

## Data Availability

All data produced in the present study are available upon reasonable request to the authors.

## Acknowledgements

Data used in the preparation of this article were obtained from the Adolescent Brain Cognitive Development^SM^ (ABCD) Study (https://abcdstudy.org), held in the NIMH Data Archive (NDA). This is a multisite, longitudinal study designed to recruit more than 10 000 children age 9–10 and follow them over 10 years into early adulthood. The ABCD Study is supported by the National Institutes of Health and additional federal partners under award numbers U01DA041022, U01DA041028, U01DA041048, U01DA041089, U01DA041106, U01DA041117, U01DA041120, U01DA041134, U01DA041148, U01DA041156, U01DA041174, U24DA041123, U24DA041147, U01DA041093, and U01DA041025. A full list of supporters is available at https://abcdstudy.org/federal-partners.html. A listing of participating sites and a complete listing of the study investigators can be found at https://abcdstudy.org/Consortium_Members.pdf. ABCD consortium investigators designed and implemented the study and/or provided data but did not necessarily participate in analysis or writing of this report. This manuscript reflects the views of the authors and may not reflect the opinions or views of the NIH or ABCD consortium investigators. The ABCD data repository grows and changes over time. The ABCD data used in this report came from doi:10.15154/1523041.

