## Supplementary Material for "Transdiagnostic Mental-Health Burden Shapes Cognition and Brain Structure in Adolescents: A Longitudinal Study"

Running Title: Transdiagnostic Mental-Health Burden in Adolescence

*corresponding author:

Pritha Sen/Franziska Knolle

Department of Diagnostic and Interventional Neuroradiology, School of Medicine and Health, Klinikum rechts der Isar, Technical University Munich, 81675 Munich, Germany

1. **Mental-health Questionnaires descriptions**

Supplementary Table 1. Overview of Mental-health Variables

2. **Mental-health burden score analysis with z-score approach**

**2.1 Methods**

Similar to the PCA approach, to evaluate the robustness of group definitions to the method of score construction, we implemented an alternative strategy using z-score summation. Mental-health variables were individually standardized (z-scored). For each participant, these standardized values were summed across all included variables to yield a composite “mental-health burden z-score,” representing cumulative symptom burden across domains.

Then, to identify distinct participant subgroups, we applied Gaussian Mixture Modeling (GMM) to the mental-health burden score using the Mclust package in R. The model selected the optimal number of components (k = 4) based on the Bayesian Information Criterion (BIC). Groups were labeled post hoc as Very Low*,* Low*,* Moderate*,* and High mental-health burden based on their score distributions.

All the analyses for the PCA approach was then similarly followed for the z-score approach. All cognitive and structural differences were computed including age, sex and site ID as covariates.

**2.2 Results**

**2.2.1. Mental-Health Variables Differences**

Forest plots revealed clear and consistent stratification across mental-health burden groups (Very Low, Low, Moderate, High) at each timepoint from baseline to 3-year follow-up. Across all panels in Supplementary Figure 1 (A–E), participants in the High mental-health burden group displayed the highest scores across nearly all symptom dimensions, particularly for internalizing symptoms such as depression, anxiety, suicidality, distress caused due to psychotic-like experiences (PLEs), and bipolar disorder symptoms. In contrast, the Very Low group consistently showed the lowest levels of psychopathology.

At baseline (Supplementary Figure 1A), the clearest separation between mental-health burden groups was observed for distress caused due to PLEs (distress), depression, bipolar disorder, generalized and social anxiety disorder, and prodromal psychosis syndrome (pps) scores. These variables showed a graded increase from the Very Low to High group, with the High group displaying particularly elevated scores. The remaining variables also contributed to the separation, albeit to a slightly lesser extent.

These group differences remained consistent in Year 1 (Supplementary Figure 1B), with distress and pps continuing to show strong differentiation across groups. Variables such as mania, bad life events and its impact, and suicidality were also notably elevated in the High group during this timepoint.

In Year 2 (Supplementary Figure 1C), the separation persisted across a broader set of variables, including depression, generalized anxiety disorder, distress caused due to PLEs, peer victimization, bad life event impact and bipolar disorder. The High group consistently exhibited higher levels across these dimensions compared to other groups.

By Year 3 (Supplementary Figure 1D), peer victimization, relational victimization, and peer aggression emerged as particularly distinguishing features of the High group. While symptoms such as bad life event impact and distress caused due to PLEs remained elevated, peer-related difficulties became increasingly prominent contributors to mental-health burden differentiation at this later stage.

By Year 4 (Supplementary Figure 1E), the High burden group continued to report elevated scores with maximum differences in peer victimization, distress caused due to PLEs, affected bad life events, peer aggression, peer relational and reputational victimization compared to lower burden groups.

These temporal patterns are further supported by the ridge density plots (Supplementary Figure 2 A–E), which visualize the top six variables contributing to group separation at each year. Notably, distress caused due to PLEs and bad life event impact consistently appear across years as top contributors, while peer-related variables become more dominant in later years.

Similar to the PCA approach, effect sizes remained substantial and stable across years (please see Supplementary Table 2).

.

Supplementary Figure 1. Forest plots showing group-wise comparisons of mental-health scores across timepoints.

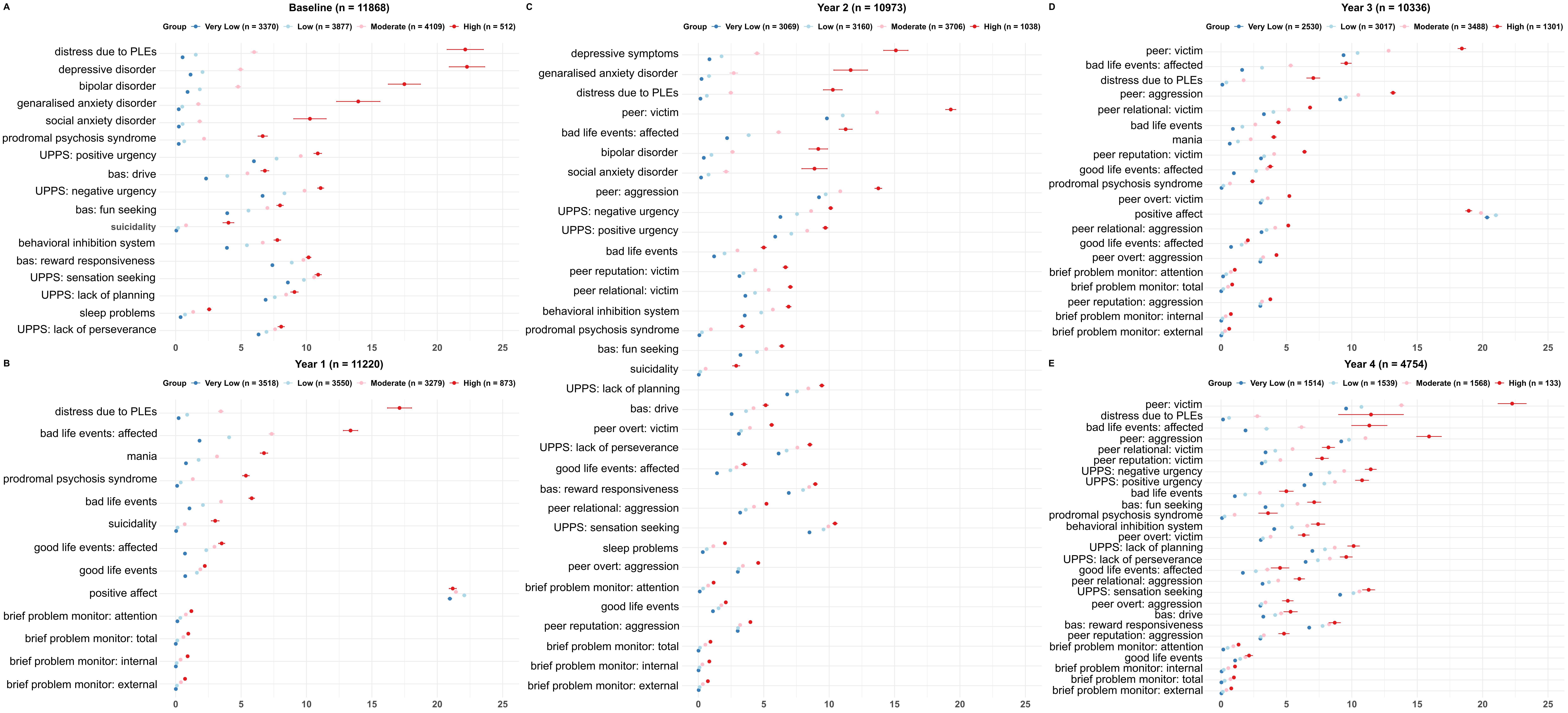

Note: Participants were grouped into four mental-health burden categories (Very Low, Low, Moderate, High) based on Gaussian Mixture Modelling applied to the first principal component of mental-health scores at each timepoint. Points reflect group means and 95% confidence intervals. Variables within each panel are ordered by effect size for the High vs. Very Low group comparison. Clear group separation is observed across all timepoints. Abbreviations: PLEs = psychotic-like experiences; UPPS = Urgency, Premeditation, Perseverance, Sensation Seeking, Positive Urgency, Impulsive Behaviour; bas = behavioural approach system. Please refer to Supplementary Table 1 for more details on the scores.

To support the rationale for deriving a composite mental-health burden score, we visualized the distribution of mental-health variables across mental-health burden groups from Baseline to 4-year follow-up. For each timepoint, we selected the top six individual symptom variables based on effect size differences between mental-health burden groups. Group-wise density plots were generated for each of these variables alongside the composite mental-health burden score. As shown in Supplementary Figure 2 (A–E), individual variables (e.g., suicidality, distress caused due to PLEs, bad life events) showed considerable distributional overlap between groups. In contrast, the composite score demonstrated clearer separation across groups (Very Low to High), highlighting its improved sensitivity and interpretability. These plots reinforce the benefit of aggregating across multiple domains to derive a more robust and continuous index of transdiagnostic mental-health burden.

Supplementary Figure 2. Distribution plots showing mental-health burden group separation, and sankey diagram showing mental-health burden group transitions over time.

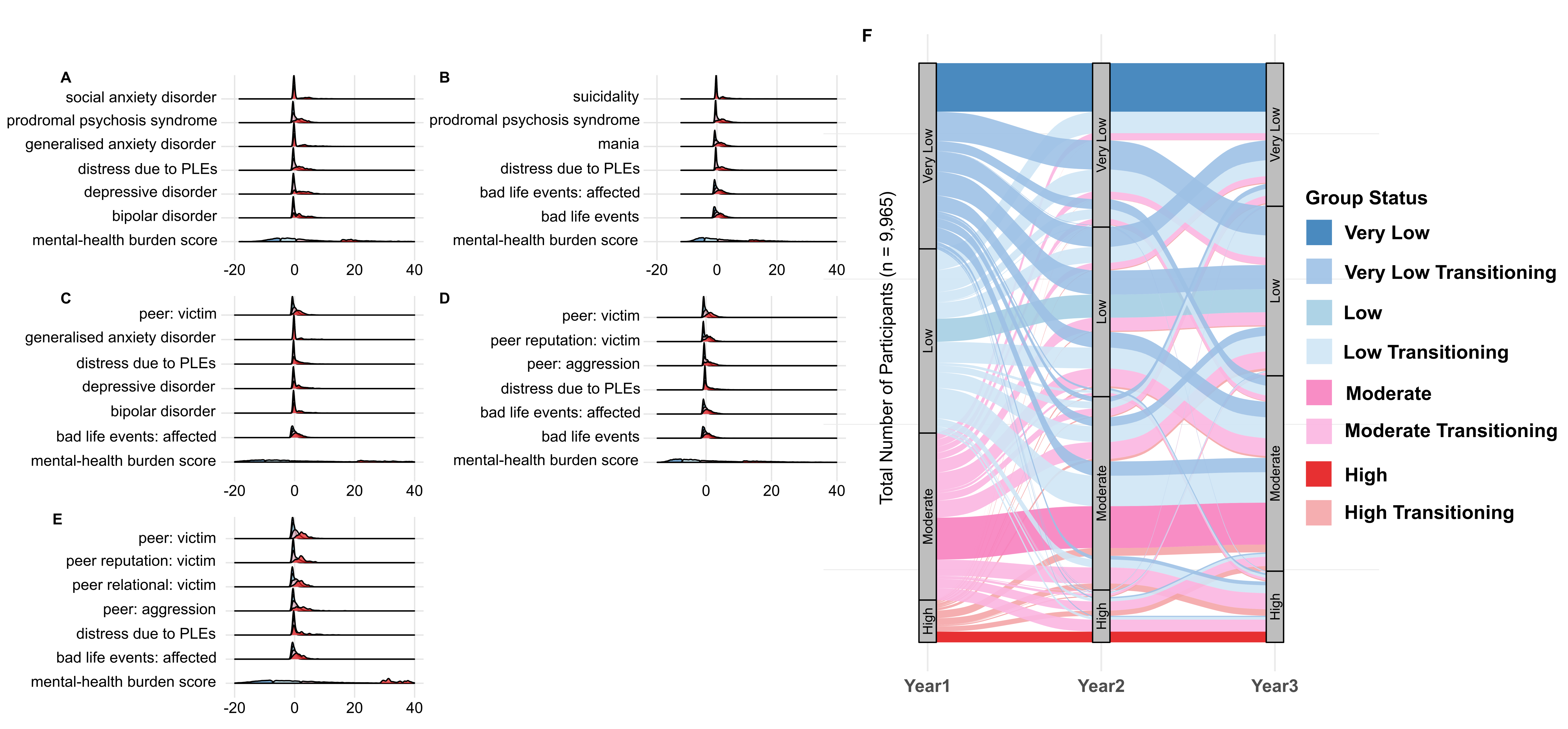

Note: (A) to (E) For each year (baseline to Year 4), the six mental-health scores shown are those with the largest absolute mean difference between the High and Very Low groups; x-axis displays the scores of the respective symptoms. Abbreviation: PLEs = psychotic-like experiences. (F) The plot displays transitions in mental-health burden group membership from 1- to 3-year follow-up among participants with complete data across all four timepoints (n=9,965). Groups were defined using z-score and Gaussian Mixture Modelling at each timepoint. Participants who remained in the same group from 1- to 3-year follow-up are labelled by their stable group (e.g., Low) and indicated in darker colours, while those who changed groups are labelled as “Transitioning” from their 1-year follow-up classification (e.g., Low Transitioning) and indicated in lighter colours. Stream widths represent the number of individuals transitioning between groups.

Supplementary Table 2. Group differences in mental-health variables based on individual year mental-health burden groups.

**2.2.2 Mental-Health Burden Group Transitions from 1- to 3-year follow-up**

To explore how participants’ mental-health burden group assignments evolved over time, we visualized transition patterns across 1-, 2- and 3-year follow-ups using a Sankey diagram (Supplementary Figure 2F). This analysis included all participants (n = 9,965) who had complete data across these four timepoints.

The width of each stream in the diagram represents the number of individuals transitioning from one group to another. Group memberships were defined separately at each timepoint using the z-score + GMM pipeline. To highlight patterns of longitudinal stability, participants whose group assignments remained consistent from Year 1 through Year 3 were labeled by their stable group. In contrast, participants whose group assignments fluctuated across years were labeled based on their Year 1 group and marked as “Transitioning”.

Similar to the PCA approach, the visualization revealed that while many participants retained their original group classification over time, there was also notable group mobility. For instance, some individuals initially in the Low group transitioned to Moderate or Very Low over subsequent years. In contrast, the High group exhibited comparatively greater stability, with fewer transitions observed.

Participants who showed stable group membership across 1-, 2- and 3- year follow-ups defined the final mental-health burden groups (Very Low: n = 835, Low: n = 397, Moderate: n = 721, High: n = 184). Demographic characteristcs for stable participants are presented in Supplementary Table 3. Further details on mental-health variable distributions for stable participants are presented in Supplementary Table 4.

Supplementary Table 3. Demographic characteristcs for stable participants with persistent mental-health burden groups from 1-year follow-up to 3-year follow-up.

Supplementary Table 4. Group differences in mental-health variables for stable participants based on persistent mental-health burden groups from 1-year follow-up to 3-year follow-up.

**2.2.3 Cognitive Performance Across Final Mental-Health Burden Groups**

We next examined whether z-score-derived mental-health burden groups differed in cognitive performance 2-year follow-up, using NIH Toolbox scores. As shown in Supplementary Figure 3, group-wise differences were observed across multiple domains, with the High mental-health burden group consistently showing poorer performance.

At 2-year follow-up (Supplementary Figure 3A), significant differences were observed across three cognitive domains including picture vocabulary (F(3, 1986) = 7.03, p < 0.001, η² = 0.01), picture sequence (F(3, 1977) = 5.78, p = 0.004, η² = 0.008), and **oral reading recognition** (F(3, 1972) = 5.33, p = 0.006, η² = 0.008). No group differences were observed for **flanker** (F(3, 1605) = 2.79, p = 0.197, η² = 0.005) or pattern comparison (F(3, 1598) = 1.38, p = 0.246, η² = 0.003) scores at Year 2.

Overall, these findings indicate that youth with persistently high mental-health burden show lower cognitive performance in domains involving language, memory, and processing speed.

All results are corrected for multiple comparisons using Bonferroni correction (p < 0.05).

Supplementary Figure 3. Group differences in cognitive performance and brain structure at baseline and 2-year follow-up, by mental-health burden group based on stable participants across 1-year follow-up through 3-year follow-up

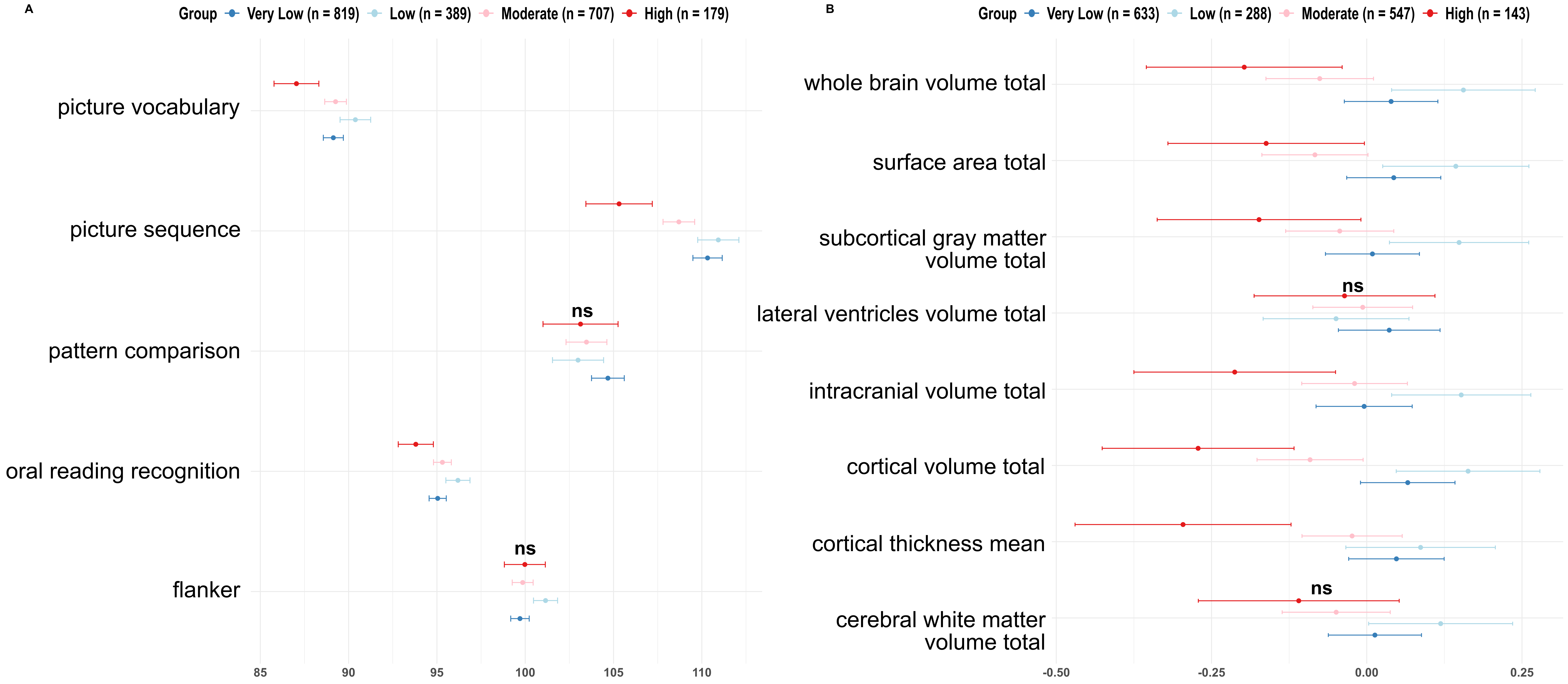

Note: (A) Cognitive tasks scores at 2-year follow-up. The High mental-health burden group scored lower across most domains. Group differences were particularly pronounced in Picture Vocabulary, Picture Sequence, and Pattern Comparison tasks. (B) Structural measures 2-year-follow-up time-point. Left and right hemisphere measures were summed to derive total values (e.g., cerebral white matter volume total =LH + RH) and subsequently standardised. Points reflect group means and 95% confidence intervals. ns indicates non-significant group differences (p<0.05) after correcting for multiple comparisons.

**2.2.4 Structural Imaging Differences Across Final Mental-Health Burden Groups**

We then examined whether mental-health burden groups differed in global structural brain measures at 2-year follow-up. At 2-year follow-up (Supplementary Figure 3B), we observed significant group differences in nearly all structural features: whole brain volume (F(3, 1607) = 7.86, p < 0.001, η² = 0.01), surface area (F(3, 1607) = 7.03, p < 0.001, η² = 0.01), subcortical gray matter volume (F(3, 1607) = 5.04, p = 0.014, η² = 0.009), intracranial volume (F(3, 1607) = 6.81, p = 0.001, η² = 0.01), cortical volume (F(3, 1607) = 11.68, p < 0.001, η² = 0.02), mean cortical thickness (F(3, 1607) = 6.09, p = 0.003, η² = 0.01). All measures except lateral ventricle volume (F(3, 1607) = 0.58, p = 1, η² = 0.001) and cerebral white matter volume (F(3, 1607) = 3.19, p = 0.183, η² = 0.006) showed significant group differences after correcting for multiple comparisons. All results are corrected for multiple comparisons using Bonferroni correction (p < 0.05).

These results suggest that youth with higher mental-health burden show widespread differences in global brain structure, consistent across both timepoints.

**2.2.5 Results for Baseline and Year 4 as Control Timepoints for Final Mental-Health Burden Groups**

To evaluate whether the mental-health burden groupings derived from stable trajectories (Year 1 to Year 3) retained predictive utility at baseline and Year 4, we examined group-level differences in mental-health variables, cognitive and structural outcomes at these time-points, using these stable group assignments. Please refer to Supplementary Table 4 for further details on mental-health score distributions and differences for baseline and 4-year follow-up as control datasets.

We observed widespread and consistent group differences in mental-health variables. All variables showed statistically significant differences across mental-health burden groups after Bonferroni correction (p < 0.05) (Supplementary Figure 1A and 1E; see Supplementary Table 4 for full ANOVA statistics). The High mental-health burden group continued to report elevated scores with topmost effect size differences in distress caused due to PLEs, affected bad life events, peer victimization and impulsivity-related measures (UPPS positive and negative urgency, fun seeking behavioral approach) compared to lower burden groups. In contrast, differences in cognitive and structural measures were no longer consistently observed at Year 4 compared to earlier time-points.

At baseline (Supplementary Figure 4A), significant group differences emerged in four of the cognitive tasks, including list sorting working memory (F(3, 2091) = 7.29, p < 0.001, η² = 0.01), dimensional change card sorting (F(3, 2101) = 4.44, p = 0.028, η² = 0.006), picture sequence (F(3, 2099) = 5.78, p = 0.004, η² = 0.008) and oral reading recognition F(3, 2098) = 3.76, p = 0.01, η² = 0.005). In contrast, no statistically significant differences were observed in flanker (F(3, 2099) = 2.77, p = 0.284, η² = 0.004), picture vocabulary (F(3, 2100) = 3.27, p = 0.142, η² = 0.005) or pattern comparison (F(3, 2094) = 2.09, p = 0.698, η² = 0.003) at baseline.

Among the NIH Toolbox cognitive measures (Supplementary Figure 4B) available at Year 4, except for Picture Vocabulary test (F(3, 942) = 4.15, p = 0.037, η² = 0.01), no other significant group differences were observed after Bonferroni correction (p < 0.05): Flanker (F(3, 661) = 1.68, p = 1, η² = 0.008), List Sorting Working Memory (F(3, 937) = 2.84, p = 0.223, η² = 0.009), Pattern Comparison (F(3, 665) = 0.12, p = 1, η² = 0.0006), Picture Sequence (F(3, 938) = 1.78, p = 0.901, η² = 0.006), Oral Reading Recognition (F(3, 936) = 2.88, p = 0.211, η² = 0.009).

At baseline (Supplementary Figure 4C), we observed significant group differences in nearly all structural features, including whole brain volume (F(3, 2103) = 8.34, p < 0.001, η² = 0.001), surface area (F(3, 2103) = 7.41, p < 0.001, η² = 0.01), subcortical gray matter volume (F(3, 2103) = 7.42, p < 0.001, η² = 0.01), intracranial volume (F(3, 2103) = 8.44, p < 0.001, η² = 0.01), cortical volume (F(3, 2103) = 10.44, p < 0.001, η² = 0.01), mean cortical thickness (F(3, 2103) = 5.12, p = 0.013, η² = 0.007), and cerebral white matter volume (F(3, 2103) = 4.28, p = 0.041, η² = 0.006). No significant differences were observed in lateral ventricle volume (F(3, 2324) = 0.8, p = 1, η² = 0.001).

At Year 4, none of the global structural brain measures (Supplementary Figure 4D), other than total cortical volume (F(3, 605) = 5.47, p = 0.008, η² = 0.03), showed significant differences across mental-health burden groups after correction for multiple comparisons: whole brain volume (F(3, 605) = 3.23, p = 0.177, η² = 0.02), surface area (F(3, 605) = 3.49, p = 0.125, η² = 0.02), subcortical grey matter volume (F(3, 605) =2.51, p = 0.465, η² = 0.01), lateral ventricle volume (F(3, 605) = 0.01, p = 1, η² = 0.00007), intracranial volume (F(3, 675) = 1.91, p = 1, η² = 0.009), cortical thickness (F(3, 605) =1.65, p = 1, η² = 0.008), cerebral white matter volume (F(3, 605) = 1.66, p = 1, η² = 0.008)).

Similar to the PCA approach, this suggests that while group differences in brain structure were observable at earlier timepoints (baseline and Year 2), these distinctions may not persist or may be less detectable at later stages, especially given the reduced sample size and partial data availability at Year 4. However, mental-health differences across groups remained robust, suggesting that the mental-health burden grouping continues to capture relevant clinical variation even at later stages.

Supplementary Figure 4. Group differences in cognitive performance and structural measures at control timepoints: baseline and 4-year follow-up, by mental-health burden group based on stable participants across 1-year follow-up through 3-year follow-up.

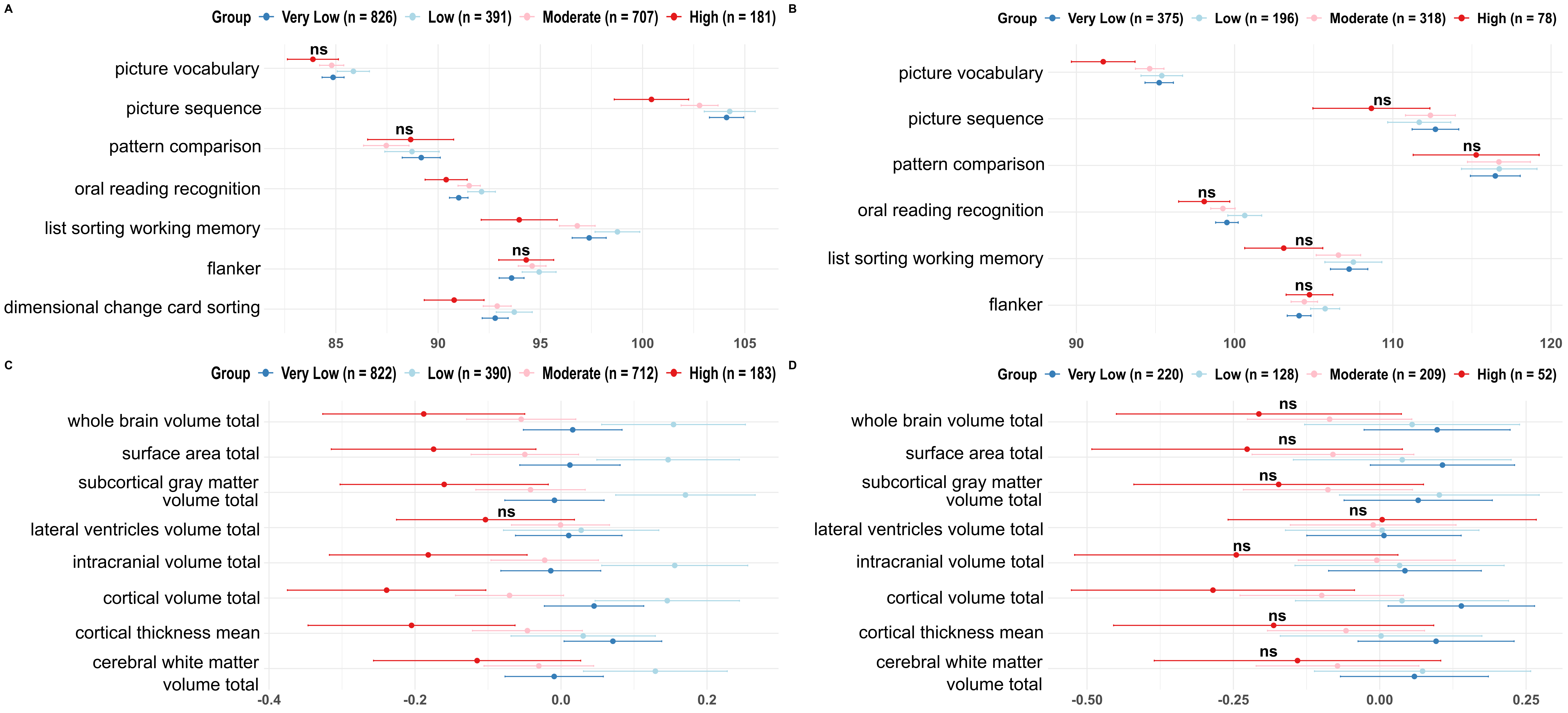

Note: Upper panel – cognitive tasks scores at (A) baseline, (B) 4-year follow-up. Lower panel – structural measures at (C) baseline and (D) 4-year-follow-up time-point. Left and right hemisphere measures were summed to derive total values (e.g., cerebral white matter volume total =LH + RH) and subsequently standardised. Points reflect group means and 95% confidence intervals. ns indicates non-significant group differences (p<0.05) after correcting for multiple comparisons.

**2.2.6 Group Differences in Cognitive Performance Based on Individual-Year Mental-Health Burden Groups**

In contrast to the previous analyses based on stable mental-health burden trajectories, the results below present cross-sectional analyses in which mental-health burden groups are defined independently at each timepoint.

At baseline (Supplementary Figure 5A), significant group-level differences were observed for all cognitive tasks, including **picture** vocabulary (F(3, 11716) = 75.09, p < 0.001, η² = 0.02), flanker (F(3, 11710) = 7.03, p < 0.001, η² = 0.018), picture sequence (F(3, 11704) = 40.01, p < 0.001, η² = 0.01), pattern comparison (F(3, 11692) = 41.34, p < 0.001, η² = 0.01), list sorting working memory (F(3, 11667) = 55.81, p < 0.001, η² = 0.01), dimensional change card sorting (F(3, 11711) = 59.02, p < 0.001, η² = 0.01) and oral reading recognition (F(3, 11702) = 49.98, p < 0.001, η² = 0.01).

Similarly, at Year 2 (Supplementary Figure 5B), group-level differences were observed for all cognitive tasks, including **picture** vocabulary (F(3, 10336) = 12.86, p < 0.001, η² = 0.004), flanker (F(3, 8181) = 5.43, p = 0.005, η² = 0.002), picture sequence (F(3, 10394) = 25.32, p < 0.001, η² = 0.007), pattern comparison (F(3, 8142) = 7.98, p < 0.001, η² = 0.003) and oral reading recognition (F(3, 10324) = 7.25, p < 0.001, η² = 0.002).

At Year 4 (Supplementary Figure 5C), after correcting for multiple comparisons, significant differences were observed only for oral reading recognition (F(3, 4532) = 5.31, p = 0.007, η² = 0.003). The other scores, including **picture** vocabulary (F(3, 4562) = 3.9, p = 0.052, η² = 0.003), flanker (F(3, 3264) = 1.29, p = 1, η² = 0.001), picture sequence (F(3, 4534) = 1.48, p = 1, η² = 0.001), pattern comparison (F(3, 3260) = 1.69, p = 1, η² = 0.002) and list sorting working memory (F(3, 4535) = 2.52, p = 0.339, η² = 0.002), remained non-significant.

All results are corrected for multiple comparisons using Bonferroni correction (p < 0.05).

Taken together, these results, yielding similar findings to the PCA approach, show that mental-health burden groups defined at individual timepoints are associated with widespread cognitive performance differences, particularly at earlier timepoints. At baseline and 2-year follow-up, the High mental-health burden group consistently showed lower scores across multiple cognitive domains, especially in picture vocabulary, working memory, and pattern comparison tasks, aligning with effect size gradients visible in Figure 7A–B. However, by Year 4, most group differences were no longer statistically significant, suggesting a potential convergence of cognitive scores over time or greater inter-individual variability in later adolescence. The only exception was oral reading recognition, which remained significantly different across groups, albeit with small effect size.

**2.2.7 Group Differences in Brain Structure Based on Individual-Year Mental-Health Burden Groups**

In this section, we examined structural brain differences using the cross-sectional mental-health burden groups defined independently at each timepoint.

Structural MRI measures also revealed significant group-level differences at both timepoints when using year-wise mental-health burden groups. At baseline, year 2 and year 4, almost all structural measures, except for lateral ventricle volume, showed significant variation across groups, showed significant variation across groups, with higher mental-health burden linked to reduced whole-brain, cortical, and white matter volumes, among others.

Baseline (Supplementary Figure 5D): whole brain volume (F(3, 11724) = 9.17, p < 0.001, η² = 0.002), surface area (F(3, 11724) = 6.49, p = 0.002, η² = 0.002), subcortical gray matter volume (F(3, 11724) = 9.38, p < 0.001, η² = 0.002), lateral ventricle volume (F(3, 11724) = 0.82, p = 1, η² = 0.0002), intracranial volume (F(3, 11724) = 10.88, p < 0.001, η² = 0.003), cortical volume (F(3, 11724) = 12.16, p < 0.001, η² = 0.003), cortical thickness (F(3, 11724) = 8.23, p < 0.001, η² = 0.002), cerebral white matter volume (F(3, 11724) = 4.4, p = 0.034, η² = 0.001).

Year 2 (Supplementary Figure 5E): whole brain volume (F(3, 8088) = 18.71, p < 0.001, η² = 0.007), surface area (F(3, 8088) = 15.61, p < 0.001, η² = 0.006), subcortical gray matter volume (F(3, 8088) = 12.73, p < 0.001, η² = 0.005), lateral ventricle volume (F(3, 8088) = 0.58, p = 1, η² = 0.0002), intracranial volume (F(3, 8088) = 11.11, p < 0.001, η² = 0.004), cortical volume (F(3, 8088) = 23.1, p < 0.001, η² = 0.008), cortical thickness (F(3, 8088) = 9.39, p < 0.001, η² = 0.003), cerebral white matter volume (F(3, 8088) = 9.68, p < 0.001, η² = 0.004).

Year 4 (Supplementary Figure 6F): whole brain volume (F(3, 3030) = 12.21, p < 0.001, η² = 0.01), surface area (F(3, 3030) = 10.63, p < 0.001, η² = 0.01), subcortical gray matter volume (F(3, 3030) = 8.73, p < 0.001, η² = 0.009), lateral ventricle volume (F(3, 3030) = 2.83, p = 0.296, η² = 0.003), intracranial volume (F(3, 3030) = 8.57, p < 0.001, η² = 0.008), cortical volume (F(3, 3030) = 11, p < 0.001, η² = 0.01), cortical thickness (F(3, 3030) = 0.65, p = 1, η² = 0.0006), cerebral white matter volume (F(3, 3030) = 8.11, p < 0.001, η² = 0.008).

All results are corrected for multiple comparisons using Bonferroni correction (p < 0.05).

The pattern of structural differences mirrored cognitive findings, with the most pronounced and consistent effects observed at baseline and 2-year follow-up. Across both timepoints, individuals in the High mental-health burden group showed lower global brain metrics, as shown in Figure 7D–E. These effects remained statistically robust after correction, with small but consistent effect sizes. Notably, lateral ventricle volume showed no group differences at any timepoint. At Year 4 (Figure 7F), significant group differences persisted across brain volume metrics significantly different in previous years, but the magnitude of these effects appeared attenuated compared to earlier years. This attenuation may reflect developmental compensation, increasing heterogeneity, or reduced power due to smaller sample size at this timepoint.

Supplementary Figure 5. Group differences in cognitive performance and brain structure at baseline, 2-year follow-up and 4-year follow-up, based on individual year mental-health burden groups.

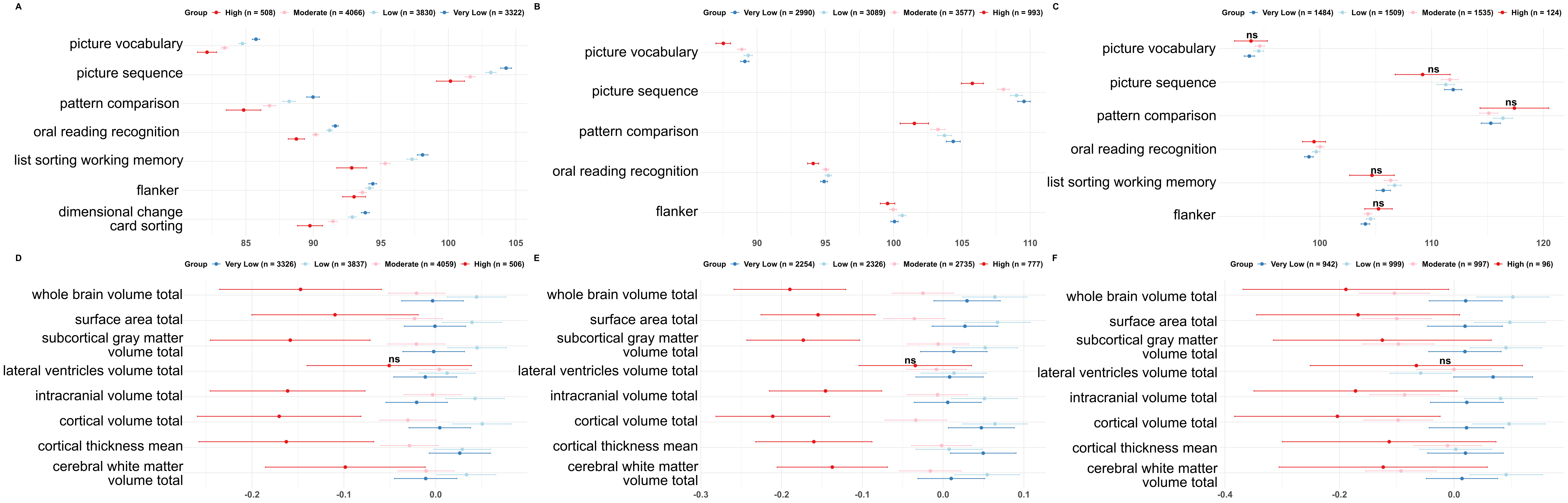

Note: Upper panel – cognitive tasks scores at (A) baseline, (B) 2-year follow-up and (C) 4-year follow-up; Lower panel – structural measures at (D) baseline, (E) 2-year follow-up and (F) 4-year follow-up. Left and right hemisphere measures were summed to derive total values (e.g., cerebral white matter volume total =LH + RH) and subsequently standardised. Points reflect group means and 95% confidence intervals. ns indicates non-significant group differences (p<0.05) after correcting for multiple comparisons.

**Mental-health burden score analysis PCA approach (including first 3 PCs)**

**3.1 Methods**

As a sensitivity analysis, we assessed whether the definition of participant groups was robust to the dimensionality of the PCA representation. Instead of applying Gaussian Mixture Modeling (GMM) to the first principal component (PC1) alone, GMM was applied to the participant scores on the first three principal components, which together explained 46.1% (baseline), 58.0% (year 1), 38.8% (year 2), 51.3% (year 3), and 42.6% (year 4) of the total variance. GMM was implemented using the mclust package in R, with the optimal number of components selected using the Bayesian Information Criterion (BIC). To facilitate comparison with the primary analyses, the resulting clusters were ordered according to their overall burden and labelled post hoc as Very Low, Low, Moderate, and High Mental-Health Burden. All subsequent behavioural, neuroimaging, and stability analyses were then repeated using these alternative group assignments.

All cognitive and structural differences were computed including age, sex and site ID as covariates.

**3.2 Results**

**3.2.1. Mental-Health Variables Differences**

Forest plots revealed clear and consistent stratification across mental-health burden groups (Very Low, Low, Moderate, High) at each timepoint from baseline to 3-year follow-up. Across all panels in Supplementary Figure 6 (A–E), participants in the High mental-health burden group displayed the highest scores across nearly all symptom dimensions, particularly for internalizing symptoms such as depression, anxiety, suicidality, distress caused due to psychotic-like experiences (PLEs), and bipolar disorder symptoms. In contrast, the Very Low group consistently showed the lowest levels of psychopathology.

At baseline (Supplementary Figure 6A), the clearest separation between mental-health burden groups was observed for distress caused due to PLEs (distress), depression, bipolar disorder, generalized and social anxiety disorder, and prodromal psychosis syndrome (pps) scores. These variables showed a graded increase from the Very Low to High group, with the High group displaying particularly elevated scores. The remaining variables also contributed to the separation, albeit to a slightly lesser extent.

These group differences remained consistent in Year 1 (Supplementary Figure 6B), with distress continuing to show strong differentiation across groups. Variables such as mania, positive affect, bad life events and its impact as well as good life events impact were also notably elevated in the High group during this timepoint.

In Year 2 (Supplementary Figure 6C), the separation persisted across a broader set of variables, including depression, social and generalized anxiety disorder, distress caused due to PLEs, peer victimization and bad life event impact. The High group consistently exhibited higher levels across these dimensions compared to other groups.

By Year 3 (Supplementary Figure 6D), peer victimization, relational and reputational victimization, and peer aggression emerged as particularly distinguishing features of the High group. While symptoms such as bad life event impact and distress caused due to PLEs remained elevated, peer-related difficulties became increasingly prominent contributors to mental-health burden differentiation at this later stage.

By Year 4 (Supplementary Figure 6E), the High burden group continued to report elevated scores with maximum differences in peer victimization, distress caused due to PLEs, affected bad life events, peer aggression, peer relational and reputational victimization compared to lower burden groups.

These temporal patterns are further supported by the ridge density plots (Supplementary Figure 7A–E), which visualize the top six variables contributing to group separation at each year. Notably, distress caused due to PLEs and bad life event impact consistently appear across years as top contributors, while peer-related variables become more dominant in later years.

Supplementary Figure 6. Forest plots showing group-wise comparisons of mental-health scores across timepoints.

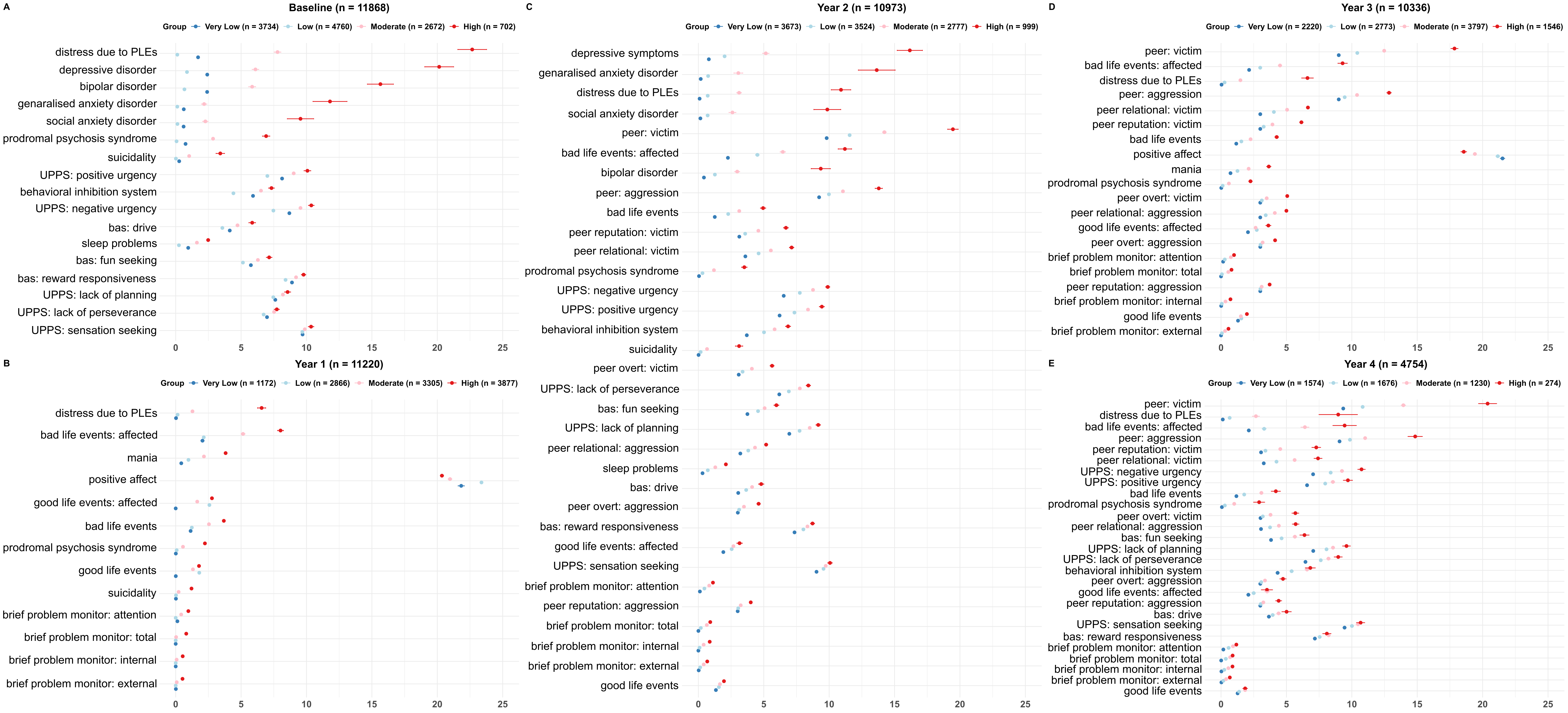

Note: Participants were grouped into four mental-health burden categories (Very Low, Low, Moderate, High) based on Gaussian Mixture Modelling applied to the first principal component of mental-health scores at each timepoint. Points reflect group means and 95% confidence intervals. Variables within each panel are ordered by effect size for the High vs. Very Low group comparison. Clear group separation is observed across all timepoints. Abbreviations: PLEs = psychotic-like experiences; UPPS = Urgency, Premeditation, Perseverance, Sensation Seeking, Positive Urgency, Impulsive Behaviour; bas = behavioural approach system. Please refer to Supplementary Table 1 for more details on the scores.

To support the rationale for deriving a composite mental-health burden score, we visualized the distribution of mental-health variables across mental-health burden groups from Baseline to 4-year follow-up. For each timepoint, we selected the top six individual symptom variables based on effect size differences between mental-health burden groups. Group-wise density plots were generated for each of these variables alongside the composite mental-health burden score. As shown in Supplementary Figure 7(A–E), individual variables (e.g., distress caused due to PLEs, bad life events) showed considerable distributional overlap between groups. In contrast as in the PCA and z-score based approaches, the composite score demonstrated clearer separation across groups (Very Low to High), highlighting its improved sensitivity and interpretability. These plots reinforce the benefit of aggregating across multiple domains to derive a more robust and continuous index of transdiagnostic mental-health burden.

Supplementary Figure 7. Distribution plots showing mental-health burden group separation, and sankey diagram showing mental-health burden group transitions over time.

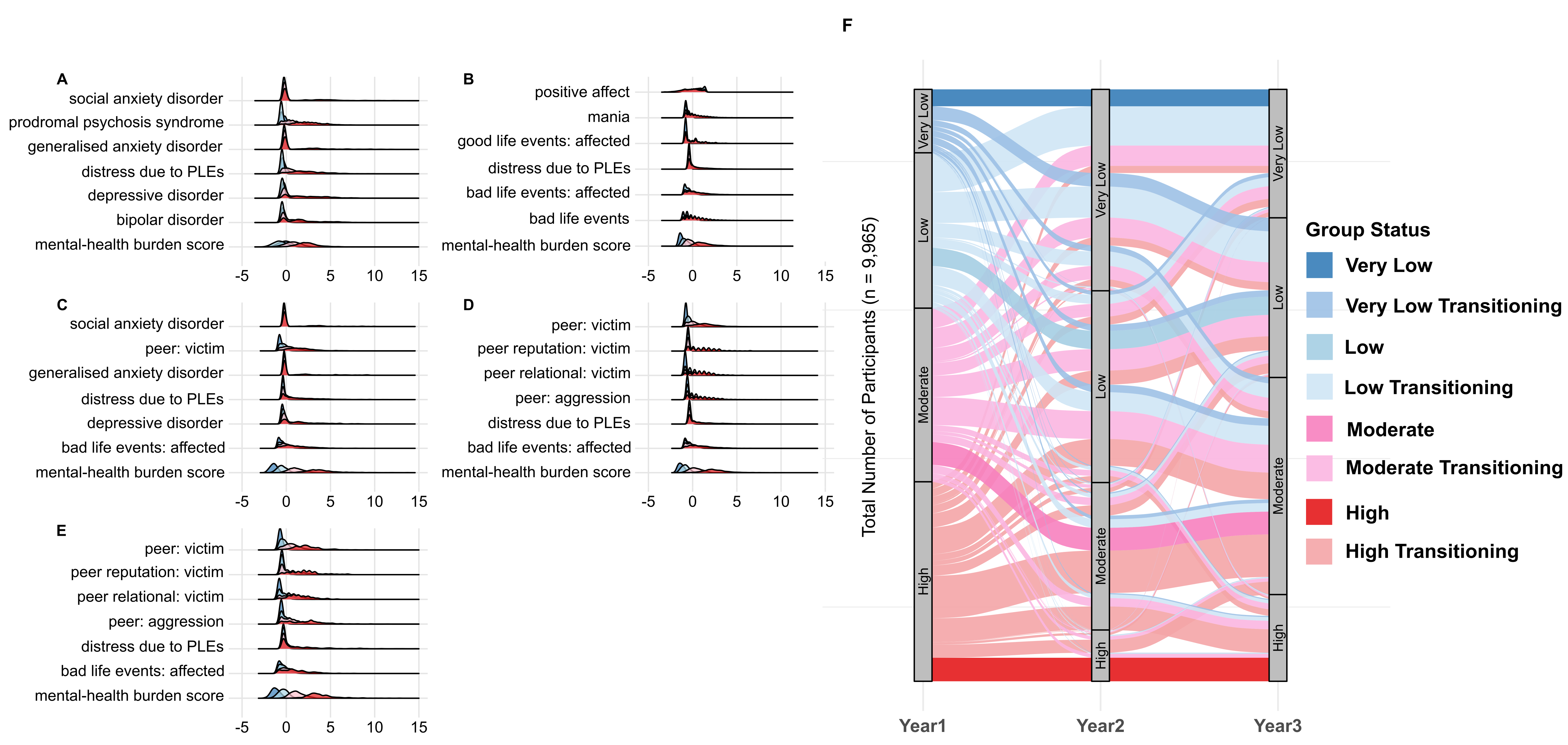

Note: (A) to (E) For each year (baseline to Year 4), the six mental-health scores shown are those with the largest absolute mean difference between the High and Very Low groups; x-axis displays the scores of the respective symptoms. Abbreviation: PLEs = psychotic-like experiences. (F) The plot displays transitions in mental-health burden group membership from 1- to 3-year follow-up among participants with complete data across all four timepoints (n=9,965). Groups were defined using PCA (first 3 PCs) and Gaussian Mixture Modelling at each timepoint. Participants who remained in the same group from 1- to 3-year follow-up are labelled by their stable group (e.g., Low) and indicated in darker colours, while those who changed groups are labelled as “Transitioning” from their 1-year follow-up classification (e.g., Low Transitioning) and indicated in lighter colours. Stream widths represent the number of individuals transitioning between groups.

**3.2.2 Mental-Health Burden Group Transitions from 1- to 3-year follow-up**

To explore how participants’ mental-health burden group assignments evolved over time, we visualized transition patterns across 1-, 2- and 3-year follow-ups using a Sankey diagram (Supplementary Figure 7F). This analysis included all participants (n = 9,965) who had complete data across these four timepoints.

The width of each stream in the diagram represents the number of individuals transitioning from one group to another. Group memberships were defined separately at each timepoint using the First 3PCs + GMM pipeline. To highlight patterns of longitudinal stability, participants whose group assignments remained consistent from Year 1 through Year 3 were labeled by their stable group. In contrast, participants whose group assignments fluctuated across years were labeled based on their Year 1 group and marked as “Transitioning”.

The visualization revealed that while many participants retained their original group classification over time, there was also notable group mobility. For instance, some individuals initially in the Low group transitioned to Moderate or Very Low over subsequent years. Compared to the PCA and z-score approach, the High group showed more movement within the groups from their starting point.

Participants who showed stable group membership across 1-, 2- and 3- year follow-ups defined the final mental-health burden groups (Very Low: n = 285, Low: n = 308, Moderate: n = 384, High: n = 396).

**3.2.3 Cognitive Performance Across Final Mental-Health Burden Groups**

We next examined whether 3PC-derived mental-health burden groups differed in cognitive performance at Year 2, using NIH Toolbox scores. As shown in Supplementary Figure 8, group-wise differences were observed across multiple domains, with the High mental-health burden group consistently showing poorer performance.

At 2-year follow-up (Supplementary Figure 8A), significant differences were observed across four cognitive domains including picture vocabulary (F(3, 1305) = 17.14, p < 0.001, η² = 0.04), picture sequence (F(3, 1304) = 11.42, p < 0.001, η² = 0.03), **flanker** (F(3, 1051) = 4.28, p = 0.026, η² = 0.01) and **oral reading recognition** (F(3, 1302) = 10.13, p < 0.006, η² = 0.02). No group differences were observed for pattern comparison (F(3, 1046) = 2.91, p = 0.168, η² = 0.008) scores at Year 2.

Overall, these findings indicate that youth with persistently high mental-health burden show lower cognitive performance in domains involving language, memory, and processing speed.

All results are corrected for multiple comparisons using Bonferroni correction (p < 0.05).

Supplementary Figure 8. Group differences in cognitive performance and brain structure at baseline and 2-year follow-up, by mental-health burden group based on stable participants across 1-year follow-up through 3-year follow-up

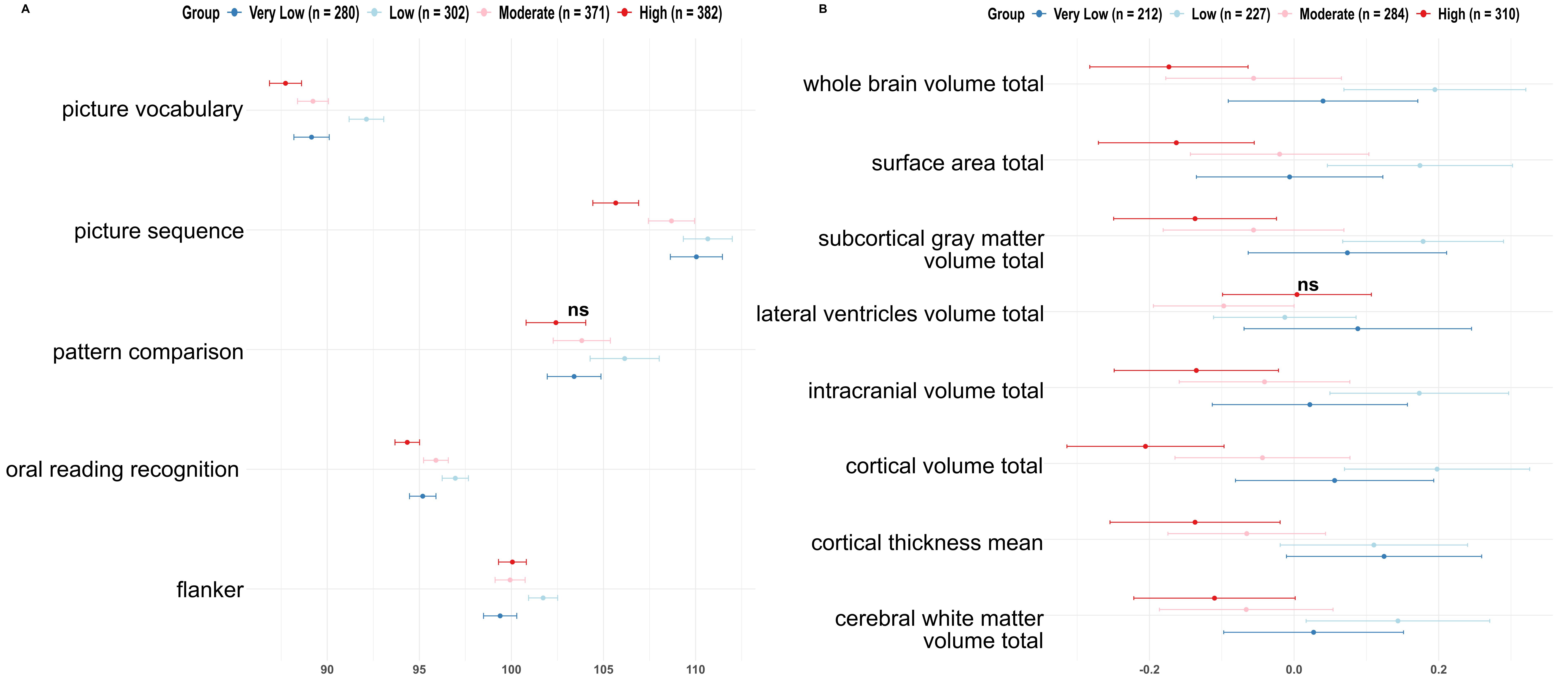

Note: (A) Cognitive tasks scores at 2-year follow-up. The High mental-health burden group scored lower across most domains. Group differences were particularly pronounced in Picture Vocabulary, Picture Sequence, and Pattern Comparison tasks. (B) Structural measures 2-year-follow-up time-point. Left and right hemisphere measures were summed to derive total values (e.g., cerebral white matter volume total =LH + RH) and subsequently standardised. Points reflect group means and 95% confidence intervals. ns indicates non-significant group differences (p<0.05) after correcting for multiple comparisons.

**3.2.4 Structural Imaging Differences Across Final Mental-Health Burden Groups**

We then examined whether mental-health burden groups differed in global structural brain measures at 2-year follow-up. At 2-year follow-up (Supplementary Figure 8B), we observed significant group differences in nearly all structural features, including whole brain volume (F(3, 1029) = 8.99, p < 0.001, η² = 0.03), surface area (F(3, 1029) = 7.16, p < 0.001, η² = 0.02), subcortical gray matter volume (F(3, 1029) = 6.4, p = 0.002, η² = 0.02), intracranial volume (F(3, 1029) = 6.27, p = 0.003, η² = 0.02), cortical volume (F(3, 1029) = 10.52, p < 0.001, η² = 0.03), mean cortical thickness (F(3, 1029) = 4.7, p = 0.023, η² = 0.01) and cerebral white matter volume (F(3, 1029) = 4.3, p = 0.041, η² = 0.01). All measures except lateral ventricle volume (F(3, 1029) = 1.68, p = 1, η² = 0.005) showed significant group differences after correcting for multiple comparisons. All results are corrected for multiple comparisons using Bonferroni correction (p < 0.05).

These results suggest that youth with higher mental-health burden show widespread differences in global brain structure.

**3.2.5 Results for Baseline and Year 4 as Control Timepoints for Final Mental-Health Burden Groups**

To evaluate whether the mental-health burden groupings derived from stable trajectories (Year 1 to Year 3) retained predictive utility at baseline and Year 4, we examined group-level differences in mental-health variables, cognitive and structural outcomes at these time-points, using these stable group assignments.

We observed widespread and consistent group differences in mental-health variables. All variables showed statistically significant differences across mental-health burden groups after Bonferroni correction (p < 0.05) (Supplementary Figure 6A and 6E). The High mental-health burden group continued to report elevated scores with topmost effect size differences in distress caused due to PLEs, depression, anxiety, affected bad life events and peer victimization compared to lower burden groups. In contrast, differences in cognitive and structural measures were no longer consistently observed at Year 4 compared to earlier time-points.

At baseline (Supplementary Figure 9A), significant group differences emerged in five of the cognitive tasks, including list sorting working memory (F(3, 1350) = 13.21, p < 0.001, η² = 0.03), dimensional change card sorting (F(3, 1354) = 9.82, p < 0.001, η² = 0.02) and picture sequence (F(3, 1351) = 9.13, p < 0.001, η² = 0.02), picture vocabulary (F(3, 1354) = 8.99, p < 0.001, η² = 0.02) and oral reading recognition F(3, 1352) = 7.78, p < 0.001, η² = 0.02). No statistically significant differences were observed for flanker (F(3, 1354) = 3.79, p = 0.071, η² = 0.008) or pattern comparison (F(3, 1351) = 3.23, p = 0.153, η² = 0.007) at baseline.

Among the NIH Toolbox cognitive measures (Supplementary Figure 9B) available at Year 4, except for Picture Vocabulary test (F(3, 595) = 4.34, p = 0.029, η² = 0.02), no other significant group differences were observed after Bonferroni correction (p < 0.05): Flanker (F(3, 428) = 0.97, p = 1, η² = 0.007), List Sorting Working Memory (F(3, 593) = 3.33, p = 0.116, η² = 0.02), Pattern Comparison (F(3, 430) = 0.91, p = 1, η² = 0.007), Picture Sequence (F(3, 590) = 1.44, p = 1, η² = 0.007), Oral Reading Recognition (F(3, 592) = 2.98, p = 0.185, η² = 0.02).

At baseline (Supplementary Figure 9C), we observed significant group differences in nearly all structural features, including whole brain volume (F(3, 1362) = 9.32, p < 0.001, η² = 0.02), surface area (F(3, 1362) = 6.36, p = 0.002, η² = 0.01), subcortical gray matter volume (F(3, 1362) = 6.96, p < 0.001, η² = 0.02), intracranial volume (F(3, 1362) = 7.77, p < 0.001, η² = 0.02), cortical volume (F(3, 1362) = 10.58, p < 0.001, η² = 0.02), mean cortical thickness (F(3, 1362) = 6.25, p = 0.003, η² = 0.01), and cerebral white matter volume (F(3, 1362) = 4.44, p = 0.033, η² = 0.01). No significant differences were observed in lateral ventricle volume (F(3, 1362) = 1.06, p = 1, η² = 0.002).

At Year 4 (Supplementary Figure 9D), except for whole brain volume (F(3, 386) = 4.24, p = 0.047, η² = 0.03), total cortical volume (F(3, 386) = 5.5, p = 0.008, η² = 0.04) and cortical thickness (F(3, 386) =4.54, p = 0.031, η² = 0.04), no other global structural brain measures showed significant differences across mental-health burden groups after correction for multiple comparisons: surface area (F(3, 386) = 2.33, p = 0.59, η² = 0.02), subcortical grey matter volume (F(3, 386) =2.54, p = 0.448, η² = 0.02), lateral ventricle volume (F(3, 386) = 0.52, p = 1, η² = 0.004), intracranial volume (F(3, 386) = 2.63, p = 0.401, η² = 0.02), , cerebral white matter volume (F(3, 386) = 2.22, p = 0.687, η² = 0.02)).

Similar to PCA and z-score based grouping, this suggests that while group differences in brain structure were observable at earlier timepoints (baseline and Year 2), these distinctions may not persist or may be less detectable at later stages. However, mental-health differences across groups remained robust, suggesting that the mental-health burden grouping continues to capture relevant clinical variation even at later stages.

Supplementary Figure 9. Group differences in cognitive performance and structural measures at control timepoints: baseline and 4-year follow-up, by mental-health burden group based on stable participants across 1-year follow-up through 3-year follow-up.

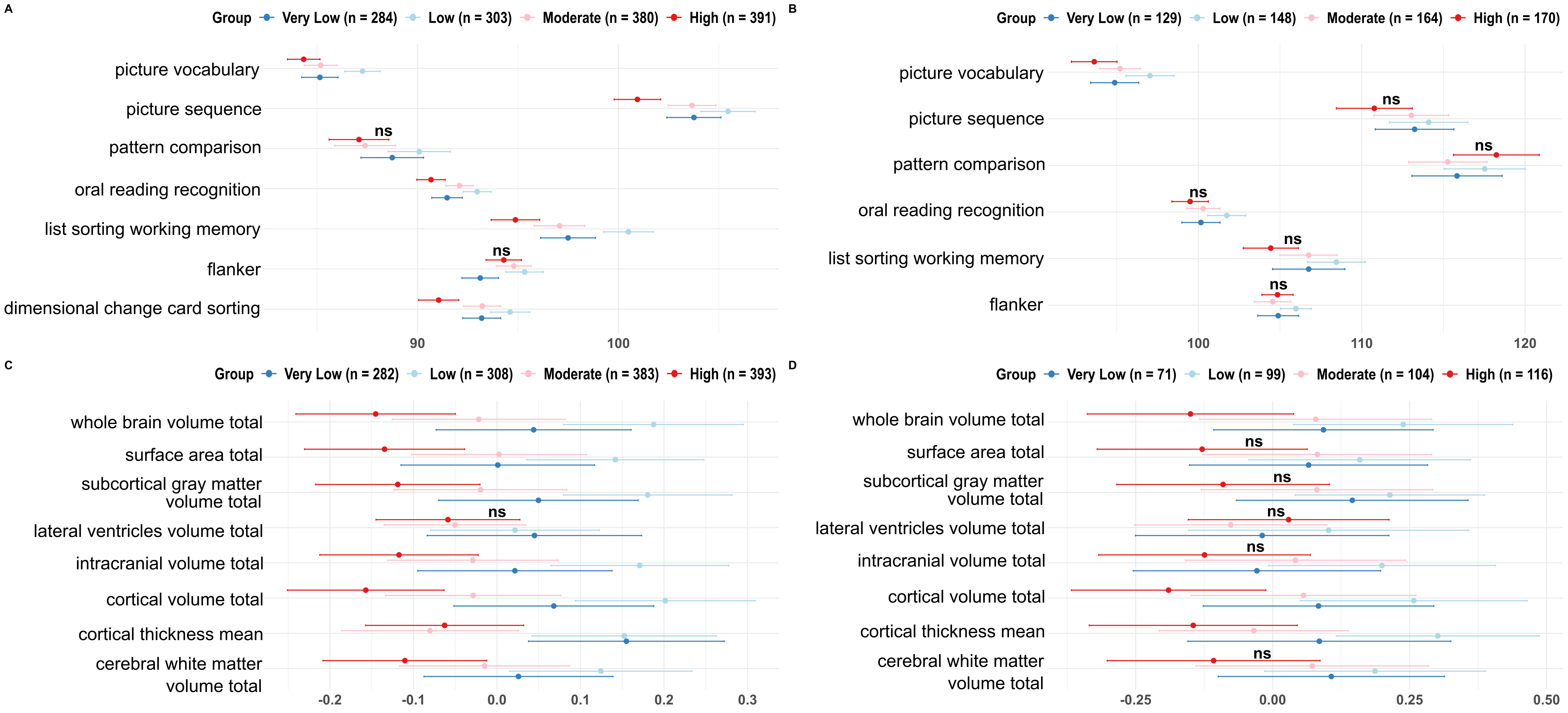

Note: Note: Upper panel – cognitive tasks scores at (A) baseline, (B) 4-year follow-up. Lower panel – structural measures at (C) baseline and (D) 4-year-follow-up time-point. Left and right hemisphere measures were summed to derive total values (e.g., cerebral white matter volume total =LH + RH) and subsequently standardised. Points reflect group means and 95% confidence intervals. ns indicates non-significant group differences (p<0.05) after correcting for multiple comparisons.

**3.2.6 Group Differences in Cognitive Performance Based on Individual-Year Mental-Health Burden Groups**

In contrast to the previous analyses based on stable mental-health burden trajectories, the results below present cross-sectional analyses in which mental-health burden groups are defined independently at each timepoint.

At baseline (Supplementary Figure 10A), significant group-level differences were observed for all cognitive tasks, including **picture** vocabulary (F(3, 11716) = 65.24, p < 0.001, η² = 0.02), flanker (F(3, 11710) = 11.17, p < 0.001, η² = 0.003), picture sequence (F(3, 11704) = 20.82, p < 0.001, η² = 0.005), pattern comparison (F(3, 11692) = 22.02, p < 0.001, η² = 0.006), list sorting working memory (F(3, 11667) = 47.93, p < 0.001, η² = 0.01), dimensional change card sorting (F(3, 11711) = 40.87, p < 0.001, η² = 0.01) and oral reading recognition (F(3, 11702) = 44.73, p < 0.001, η² = 0.01).

Similarly, at Year 2 (Supplementary Figure 10B), group-level differences were observed for all cognitive tasks, including **picture** vocabulary (F(3, 10336) = 15.09, p < 0.001, η² = 0.004), flanker (F(3, 8181) = 6.15, p = 0.002, η² = 0.002), picture sequence (F(3, 10394) = 31.83, p < 0.001, η² = 0.009), pattern comparison (F(3, 8142) = 8.72, p < 0.001, η² = 0.003) and oral reading recognition (F(3, 10324) = 7.73, p < 0.001, η² = 0.002).

At Year 4 (Supplementary Figure 10C), after correcting for multiple comparisons, significant differences were observed for oral reading recognition (F(3, 4532) = 6.38, p = 0.002, η² = 0.004) and **picture** vocabulary (F(3, 4562) = 7.72, p < 0.001, η² = 0.005). The other scores, including flanker (F(3, 3264) = 0.64, p = 1, η² = 0.0006), picture sequence (F(3, 4534) = 0.98, p = 1, η² = 0.0006), pattern comparison (F(3, 3260) = 1.73, p = 0.953, η² = 0.002) and list sorting working memory (F(3, 4535) = 2.02, p = 0.653, η² = 0.001), remained non-significant.

All results are corrected for multiple comparisons using Bonferroni correction (p < 0.05).

Similar to the PCA and z-score approaches, these results show that mental-health burden groups defined at individual timepoints are associated with widespread cognitive performance differences, particularly at earlier timepoints. At baseline and 2-year follow-up, the High mental-health burden group consistently showed lower scores across multiple cognitive domains. However, by Year 4, most group differences were no longer statistically significant, suggesting a potential convergence of cognitive scores over time or greater inter-individual variability in later adolescence. The only exceptions were oral reading recognition and picture vocabulary, which remained significantly different across groups, albeit with small effect size.

**3.2.7 Group Differences in Brain Structure Based on Individual-Year Mental-Health Burden Groups**

Structural MRI measures also revealed significant group-level differences at both timepoints when using year-wise mental-health burden groups. At baseline and year 2, almost all structural measures, except for lateral ventricle volume, showed significant variation across groups, showed significant variation across groups, with higher mental-health burden linked to reduced whole-brain, cortical, and white matter volumes, among others. At year 4, along with lateral ventricle volume, differences in cortical thickness also remained non-significant among the groups.

Baseline (Supplementary Figure 10D): whole brain volume (F(3, 11724) = 16.53, p < 0.001, η² = 0.004), surface area (F(3, 11724) = 11.84, p < 0.001, η² = 0.003), subcortical gray matter volume (F(3, 11724) = 14.89, p < 0.001, η² = 0.004), lateral ventricle volume (F(3, 11724) = 1.05, p = 1, η² = 0.0003), intracranial volume (F(3, 11724) = 19.75, p < 0.001, η² = 0.005), cortical volume (F(3, 11724) = 19.06, p < 0.001, η² = 0.005), cortical thickness (F(3, 11724) = 9.96, p < 0.001, η² = 0.003), cerebral white matter volume (F(3, 11724) = 9.26, p < 0.001, η² = 0.002).

Year 2 (Supplementary Figure 10E): whole brain volume (F(3, 8088) = 26.13, p < 0.001, η² = 0.01), surface area (F(3, 8088) = 18.69, p < 0.001, η² = 0.007), subcortical gray matter volume (F(3, 8088) = 18.27, p < 0.001, η² = 0.007), lateral ventricle volume (F(3, 8088) = 0.28, p = 1, η² = 0.0001), intracranial volume (F(3, 8088) = 15.79, p < 0.001, η² = 0.006), cortical volume (F(3, 8088) = 30.11, p < 0.001, η² = 0.01), cortical thickness (F(3, 8088) = 12.09, p < 0.001, η² = 0.004), cerebral white matter volume (F(3, 8088) = 13.94, p < 0.001, η² = 0.005).

Year 4 (Supplementary Figure 10F): whole brain volume (F(3, 3030) = 8.37, p < 0.001, η² = 0.008), surface area (F(3, 3030) = 7.8, p < 0.001, η² = 0.008), subcortical gray matter volume (F(3, 3030) = 7.74, p < 0.001, η² = 0.008), lateral ventricle volume (F(3, 3030) = 0.59, p = 1, η² = 0.0006), intracranial volume (F(3, 3030) = 7.07, p < 0.001, η² = 0.007), cortical volume (F(3, 3030) = 8.07, p < 0.001, η² = 0.008), cortical thickness (F(3, 3030) = 1.72, p = 1, η² = 0.002), cerebral white matter volume (F(3, 3030) = 5.12, p = 0.013, η² = 0.005).

All results are corrected for multiple comparisons using Bonferroni correction (p < 0.05).

The pattern of structural differences mirrored cognitive findings, with the most pronounced and consistent effects observed at baseline and 2-year follow-up. Across both timepoints, individuals in the High mental-health burden group showed lower global brain metrics, as shown in Supplementary Figure 10D–E. These effects remained statistically robust after correction, with small but consistent effect sizes. Notably, lateral ventricle volume showed no group differences at any timepoint. At Year 4 (Supplementary Figure 10F), significant group differences persisted across brain volume metrics significantly different in previous years, but the magnitude of these effects appeared attenuated compared to earlier years.

Supplementary Figure 10. Group differences in cognitive performance and brain structure at baseline, 2-year follow-up and 4-year follow-up, based on individual year mental-health burden groups.

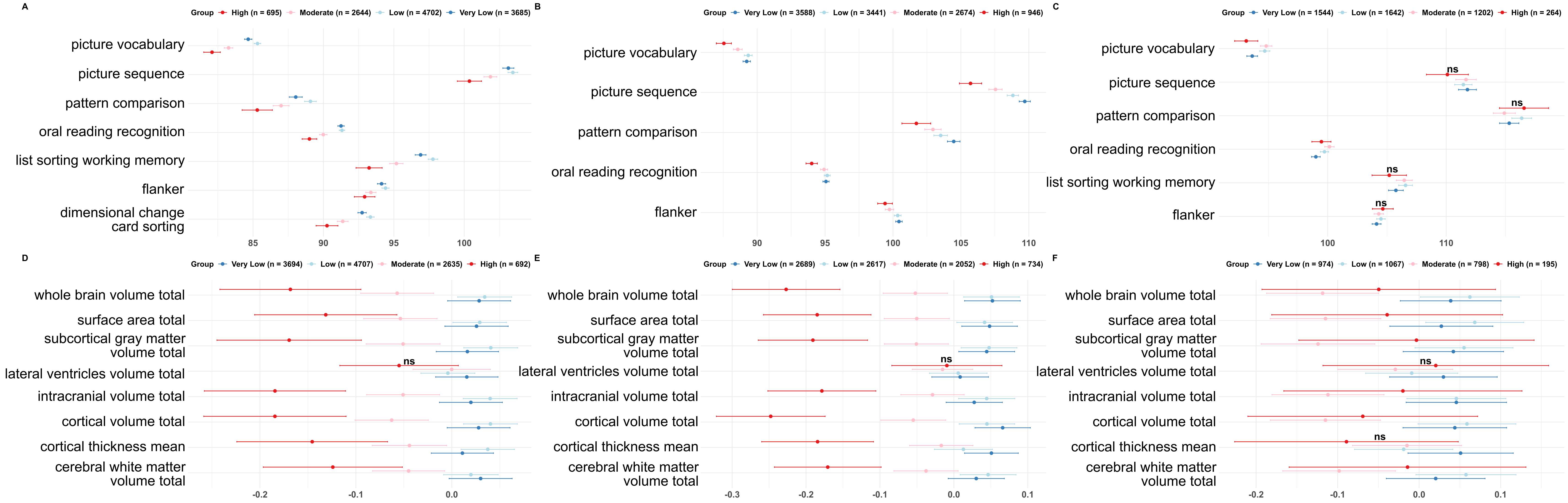

Note: Upper panel – cognitive tasks scores at (A) baseline, (B) 2-year follow-up and (C) 4-year follow-up; Lower panel – structural measures at (D) baseline, (E) 2-year follow-up and (F) 4-year follow-up. Left and right hemisphere measures were summed to derive total values (e.g., cerebral white matter volume total =LH + RH) and subsequently standardised. Points reflect group means and 95% confidence intervals. ns indicates non-significant group differences (p<0.05) after correcting for multiple comparisons.
